# Global predictions of short- to medium-term COVID-19 transmission trends : a retrospective assessment

**DOI:** 10.1101/2021.07.19.21260746

**Authors:** Sangeeta Bhatia, Kris V Parag, Jack Wardle, Natsuko Imai, Sabine L Van Elsland, Britta Lassmann, Gina Cuomo-Dannenburg, Elita Jauneikaite, H. Juliette T. Unwin, Steven Riley, Neil Ferguson, Christl A Donnelly, Anne Cori, Pierre Nouvellet

## Abstract

**Background:** As of July 2021, more than 180,000,000 cases of COVID-19 have been reported across the world, with more than 4 million deaths. Mathematical modelling and forecasting efforts have been widely used to inform policy-making and to create situational awareness.

**Methods and Findings:** From 8^th^ March to 29^th^ November 2020, we produced weekly estimates of SARS-CoV-2 transmissibility and forecasts of deaths due to COVID-19 for countries with evidence of sustained transmission. The estimates and forecasts were based on an ensemble model comprising of three models that were calibrated using only the reported number of COVID-19 cases and deaths in each country. We also developed a novel heuristic to combine weekly estimates of transmissibility and potential changes in population immunity due to infection to produce forecasts over a 4-week horizon. We evaluated the robustness of the forecasts using relative error, coverage probability, and comparisons with null models.

**Conclusions:** During the 39-week period covered by this study, we produced short- and medium-term forecasts for 81 countries. Both the short- and medium-term forecasts captured well the epidemic trajectory across different waves of COVID-19 infections with small relative errors over the forecast horizon. The model was well calibrated with 56.3% and 45.6% of the observations lying in the 50% Credible Interval in 1-week and 4-week ahead forecasts respectively. We could accurately characterise the overall phase of the epidemic up to 4-weeks ahead in 84.9% of country-days. The medium-term forecasts can be used in conjunction with the short-term forecasts of COVID-19 mortality as a useful planning tool as countries continue to relax stringent public health measures that were implemented to contain the pandemic.

## Introduction

As of July 2021, more than 4 million deaths have been attributed to COVID-19 with over 180 million cases reported globally [1]. The scale of the current pandemic has led to a widespread adoption of data-driven public health responses across the globe. Epidemiological quantities such as the reproduction number and the herd immunity threshold have become a part of the public discourse, used by governments to plan their response and by the media to aid public understanding of the health emergency. Outbreak analysis and real-time modelling, including short-term forecasts of future incidence, have been used to inform decision making and response efforts in several past public health challenges including the West African Ebola epidemic and seasonal influenza [2–11]. In the current pandemic, mathematical models have helped public health officials better understand the evolving epidemiology of SARS–CoV-2 [12–14] and the potential impact of implementing or releasing interventions. Short-term forecasts of key indicators such as mortality, hospitalisation, and hospital occupancy have played a similarly important role [15–20], contributing to planning public health interventions and allocation of crucial resources [21–25]. At the same time, the unprecedented level of public interest has placed epidemiological modelling under intense media scrutiny. While model validation against observed data is part of a typical analysis pipeline, in light of the prominent role mathematical models have had in policy planning during the COVID-19 pandemic, retrospective assessment of modelling outputs against later empirical data is critical to assess their validity.

With the aim of improving situational awareness during the ongoing pandemic, since the 8^th^ March 2020 we have been reporting weekly estimates of transmissibility of SARS-CoV-2 and forecasts of the daily incidence of deaths associated with COVID-19 for countries with evidence of sustained transmission [26]. We have developed three models that are calibrated using the latest reported incidence of COVID-19 cases and deaths in each country that we combine into an ensemble model.

All models make implicit or explicit assumptions about the data-generating process being modelled. In addition to the uncertainty associated with the model outputs, there is inherent structural uncertainty about the models themselves as each model makes different assumptions about transmission in attempting to approximate the true underlying processes. Ensemble models, which combine outputs from different models, are a powerful way of incorporating the uncertainty from a range of models [27, 28]. They are used widely in diverse fields such as weather forecasting, economics, ecology and are increasingly being used in infectious disease forecasting. Ensemble models can produce more robust forecasts than individual models [28–31]. The estimates of transmissibility and short-term forecasts presented here are based on a novel ensemble model that was calibrated across multiple waves of the pandemic.

Forecasts are typically produced under the assumption that the trend in growth remain the same over the forecast horizon. This is a plausible assumption for the 1-week forecast horizon that we used for our short-term forecasts. However, this assumption is likely to be violated over a long forecast horizon for a rapidly evolving epidemic such as COVID-19, leading to a rapidly increasing uncertainty as the forecast horizon grows. We have developed a novel approach relying on a simple heuristic that combines past estimates of the reproduction number, explicitly accounting for the predicted future changes in population immunity, to produce forecasts over longer time horizons.

Here we summarise the key transmission trends from our work on global short-term forecasts between 8^th^ March to 29^th^ November 2020. We provide a rigorous quantitative assessment of the performance of the ensemble model. We also present medium-term forecasts using our approach and retrospectively assess the performance of our method. Our results for medium-term forecasts suggest that we can accurately forecast the trajectory of COVID-19 in several countries for horizons spanning up to 4 weeks.

## Materials and methods

Methods for estimating transmissibility during epidemics typically rely on the time series of incident cases combined with the natural history parameters of the pathogen [32, 33]. However, in the current pandemic, interpretation and comparison of estimates across countries based on the number of cases was made difficult by the differences in case definitions, testing regimes, and variable reporting across countries as well as over time within each country [34]. We therefore developed methods that relied on the number of reported deaths to estimate COVID-19 transmissibility and to produce short-term forecasts of deaths. Since we use deaths to estimate transmissibility, our estimates reflect the epidemiological situation with a delay corresponding to the delay from infection to death [35]. Despite this, our estimates and the short-term forecasts contributed and continue to contribute to situational awareness by providing near real-time insights into the dynamics of the pandemic. They also provide useful, albeit indirect, evidence into the effectiveness of various interventions such as lockdowns and the impact of reopening.

The instantaneous reproduction number is frequently used to quantify transmissibility. It is defined as the average number of secondary cases that an individual infected at time *t* would generate if conditions remained as they were at time *t* [36]. When applied to the incidence of deaths (rather than cases), the instantaneous reproduction number 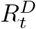 represents the average number of secondary deaths “generated by” the death of a primary case at time *t*. We developed three different models, each of which estimated transmissibility in the recent past and produced forecasts of COVID-19 deaths. We then incorporated the outputs of these models to build an ensemble model. We produced short-term forecasts (i.e. 1-week ahead), for which changes in the population immunity level could be ignored. We also produced medium-term forecasts (up to 4-weeks ahead) accounting for the depletion of the susceptible population due to the increased levels of natural host immunity. The methods underlying the individual models are illustrated in Fig. 1.

**Fig 1.**
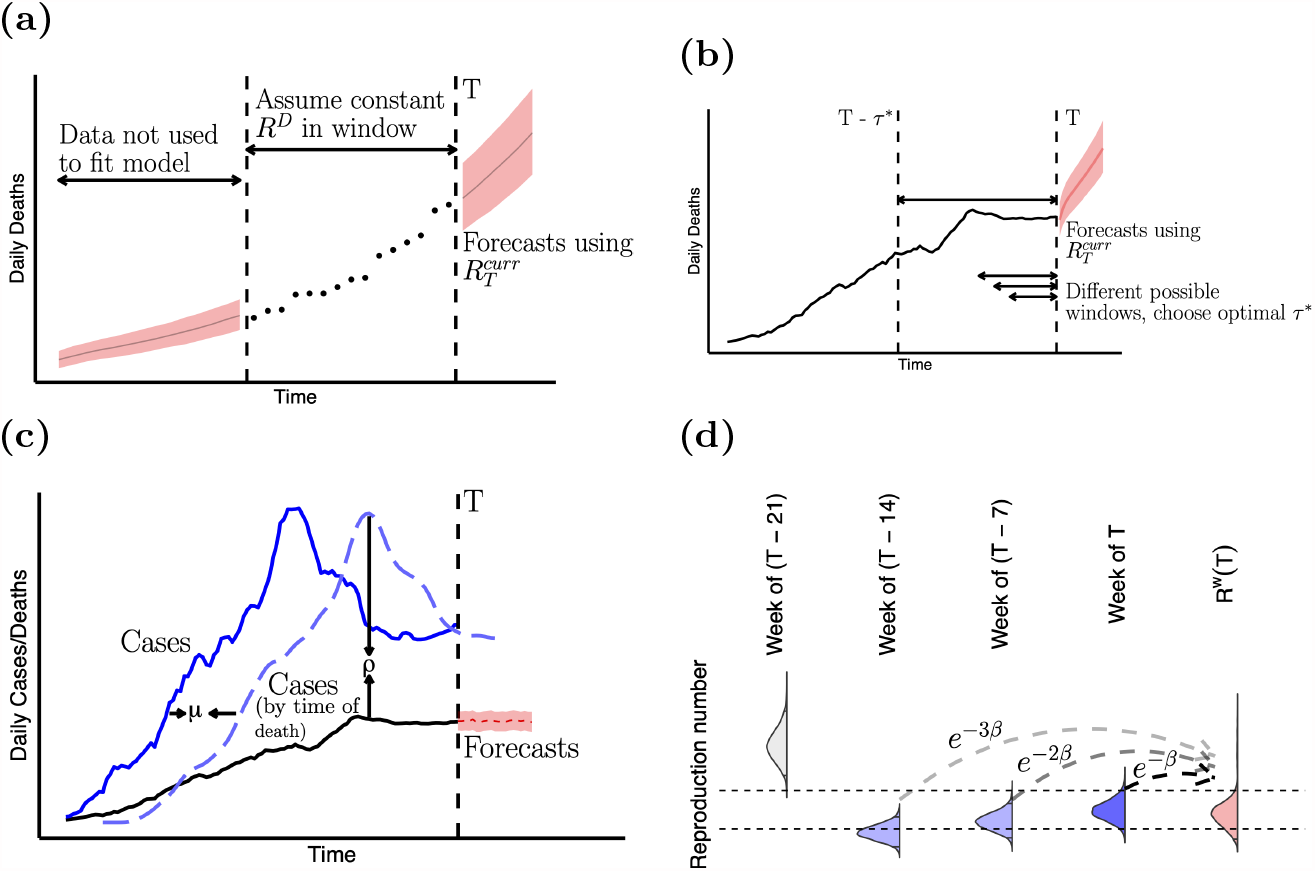
Schematic of the models. (a) Model 1 assumes a single value for *R*[*T* −*τ* +1, *T*]. The model is fitted using only the data in this window (*T* − *τ* + 1 to *T*) to jointly estimate the initial incidence of deaths and *R*[*T* − *τ* + 1, *T*]. (b) Model 2 optimises the window over which *R*_*t*_ is assumed to be constant by minimising the cumulative predictive error over the entire epidemic time series. Estimates from *R*[*T* −*τ* ^*∗*^ + 1, *T*] are used to forecast into the future, with *τ* ^*∗*^ the window of optimal length. (c) Model 3 uses data from both cases and deaths. The dashed blue curve represents the incidence of reported cases weighted by the case-report to death delay distribution, where *µ* is the mean delay. *ρ*_*t*_ is the ratio of the observed deaths and the weighted cases at time *t* and is analogous to an observed case fatality ratio. Forecasts of deaths are obtained by sampling from a binomial distribution using the most recent estimate of *ρ*_*T*_. See also SI Fig. 3. (d) To obtain medium-term forecasts, we combine the most recent transmissibility estimate 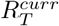 (shown in dark blue) with estimates of transmissibility in the previous weeks to produce a weighted estimate of transmissibility 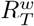 (filled in pink) at time *T*. Estimates from previous weeks are combined with the most recent estimates if the 95% CrI of estimates in week *k*, 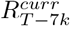 overlaps the 95% CrI of 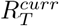. Estimates for weeks where the 95% CrI overlap are shown in light purple, and where the 95% CrI do not overlap in grey. The dashed horizontal lines represent the 2.5^th^ and 97.5^th^ quantile of 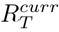. We combine the estimates by sampling from the posterior distribution of 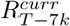 with probability proportional to *e*^*−β∗k*^, where *β* is a rate at which the probability decays as we go back in time.

Hereafter, *D*_*t*_ and *C*_*t*_ represent the number of reported COVID-19 deaths and cases at time *t* respectively. Since we only used reported deaths to estimate transmissibility, for ease of notation, we drop the superscript *D* from 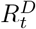 and use *R*_*t*_ to denote the instantaneous reproduction number with respect to deaths at time *t. R*[*t*_1_, *t*_2_] is the reproduction number between times *t*_1_ and *t*_2_. The most recent estimate of transmissibility is denoted as 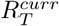. We use *ω* to denote the serial interval distribution of deaths i.e. the interval between the deaths of an infectee and their infector, where both the infector and the infectee die. Estimated incidence of deaths at time *t* is denoted by 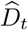. *T* refers to last time point in the existing incidence time series of cases or deaths.

### Model 1 (RtI0)

The first model relies on a well-established method [37] that assumes the daily incidence of deaths is approximated with a Poisson process following the renewal equation [36]:

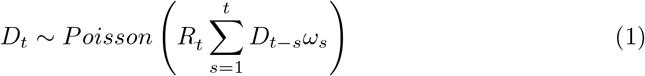

A standard approach to inferring recent transmissibility from an incidence time series relies on the assumption that the effective reproduction number is constant over a window (i.e. the “calibration window”) back in time of size *τ* time units (for example days or weeks) [33]. Adopting a similar approach here, we estimated *R*_*t*_ using only the data in a fixed time-window (of *τ* days) prior to the most recent observation to calibrate the model. We estimated the average transmissibility *R*[*T* − *τ* + 1, *T*] over that time-window, but made no assumptions regarding the epidemiological situation or transmissibility prior to this calibration window (Fig. 1a). Instead, we jointly estimated (using Markov Chain Monte Carlo (MCMC)) combinations of *R*[*T* − *τ* + 1, *T*] and the incidence of deaths prior to the calibration window 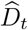 for *t* = {1, 2, … *T* − *τ*} that are consistent with the observed deaths in the time window [*T* − *τ* + 1, *T*]. The model likelihood is given by

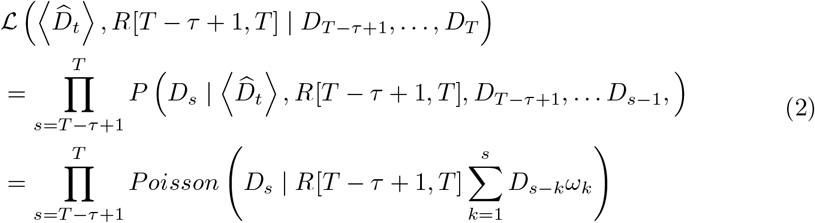

where 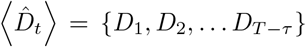 and 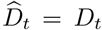 for *t* = *T* − *τ* + 1, …, *T*. The most recent estimate of transmissibility 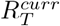 in this model is *R*[*T* − *τ* + 1, *T*]. We then sampled sets of back-calculated early incidence time series 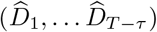 and reproduction numbers (*R*[*T* − *τ* + 1, *T*]) from the joint posterior distribution obtained in the estimation process, and projected future incidence 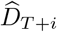 for *i* ≥ 1 conditional on these as follows:

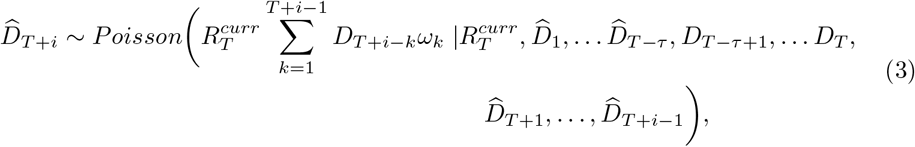

where 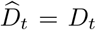 for *t* = *T* − *τ* + 1, …, *T*. During the period covered in the analysis, the epidemiological situation in most countries was changing rapidly with public health measures being reviewed weekly. At the same time, there was a strong ‘weekend effect’ in the observed data, with typically fewer deaths reported on Saturdays and Sundays. We therefore assumed a fixed calibration window of 10 days to incorporate the rapid dynamics and offset the lower reporting over the weekend. We ran the MCMC for 10000 iterations. We sampled 1000 sets of 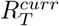 and back-calculated incidence, and for each sampled set, we drew 10 stochastic realisations of the projected incidence of deaths.

### Model 2 (APEestim)

Similarly to Model 1, Model 2 relies on the renewal equation (Eq. (1)) but uses the full time series of observed deaths, and uses information theory to optimise the choice of the calibration window i.e. the time-window of size *τ* over which *R*[*T* − *τ* + 1, *T*] is assumed to be constant in the estimation process [38]. Choices of window size can influence the bias and variance of resulting estimates of transmissibility [39]. We integrated over the entire posterior distribution of *R*_*t*_ (under a given window size), to obtain the posterior predictive distribution of incidence at time *t* + 1 as

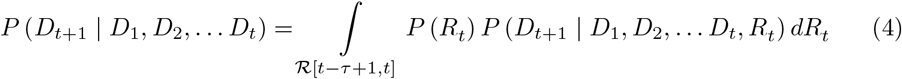

where ℛ [*t* − *τ* + 1, *t*] represents the posterior distribution of *R*_*t*_ assuming a window of length *τ*. We computed this distribution sequentially for *t* ∈ {1, 2, … *T* − 1} and then evaluated every observed count of deaths according to their likelihood under the posterior predictive distribution. This allowed us to construct the accumulated predictive error (APE) for a window length *τ* and under a given serial interval distribution as [38]:

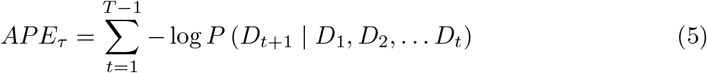

Here, *D*_*t*+1_ is the observed number of deaths at time *t*+1. The optimal window length *τ* ^*∗*^ was then chosen as the window for which *APE*_*τ*_ is minimised (Fig. 1b), optimising the bias-variance trade-off (long windows reduce the estimate variance but increase bias and short windows do the converse).

Again, forward projections were made assuming that transmissibility over the projection horizon remains the same as that in the last *τ* ^*∗*^ days. That is, 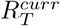 is set to be *R*[*T* − *τ* ^*∗*^ + 1, *T*]. We then obtain forecasts of deaths as

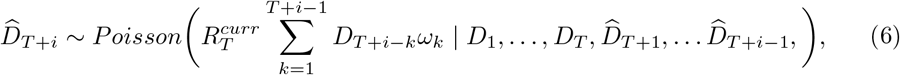

for *i* ≥ 1. We drew 1000 samples from the posterior distribution of 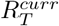 and for each sampled value, simulated 10 forward trajectories.

### Model 3 (DeCa)

Models 1 (RtI0) and 2 (APEestim) use only the time series of deaths to estimate *R*_*t*_. Model 3 exploits the signal from both the reported deaths and cases to forecast deaths. We assumed that the delay *δ* between a case being reported and the case dying (for those who die) is distributed according to a gamma distribution with mean *µ* and standard deviation *σ*. Let *f*_Γ_ be the probability mass function of a discretised gamma distribution. The cumulative number of reported cases at time *t* weighted by the delay distribution from case report to death, 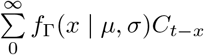, represents the potential number of deaths at time *t*, if all cases were to die. The ratio *ρ*_*t*_ of the observed number of deaths to this quantity at time *t* can be thought of as an observed case fatality ratio (Fig. 1c). We assume that deaths are distributed according to a binomial distribution:

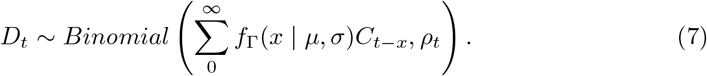

The model likelihood is given by

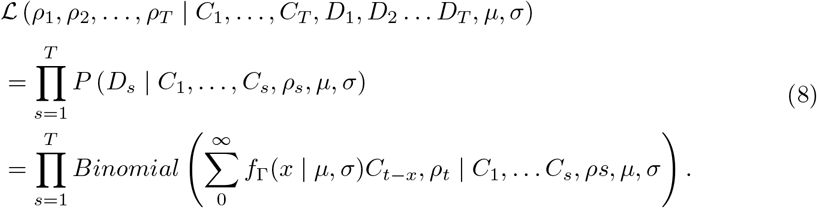

We obtained a posterior distribution for *ρ*_1_, *ρ*_2_, …, *ρ*_*T*_ using the conjugate beta prior for *ρ*_*t*_ (using the R package binom [40]), assuming that parameters of the delay distribution *µ* and *σ* are known and fixed. The forecasted number of deaths at time *T* + *i* for *i* ≥ 1 were then drawn from a binomial distribution as

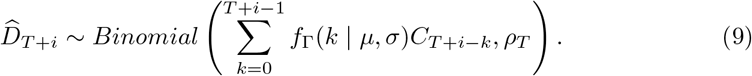

Note that the number of deaths at time *T* + *i* depends on the number of cases from the beginning of the epidemic to time *T* + *i* for *i* ≥ 1. That is, for forecasting deaths at time *T* + *i*, we need the number of cases at time *t > T*. To augment the observed time series of cases, we assumed that the cases in beyond *T* are distributed according to a gamma distribution with mean and standard deviation of the observed cases in the last week, implicitly assuming no growth or decline in cases. We assessed the extent to which this assumption affected our results (SI Sec. 4). Finally, to include transmissibility estimates from this model in the ensemble, we estimated 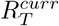 using the observed and median forecasted deaths 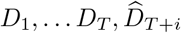 for *i* ≥ 1. using the R package EpiEstim [33].

We drew 10000 samples from the posterior distribution of *ρ*_*T*_ and 10000 samples from a gamma distribution to augment the observed cases. We then drew 10000 samples from a binomial distribution (Eq. (9)) for each pair of augmented cases trajectory and sampled *ρ*_*T*_.

### Ensemble Model

We combined the estimates of 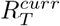, and the outputs of models RtI0, APEestim, and DeCa into an unweighted ensemble model by sampling the forecasts and reproduction number from each model described above. We also explored building a weighted ensemble by weighting the contribution of each model according to the relative error of the model in the previous week, all previous weeks, across all countries, or estimating the weights independently for each country. We did not find any substantial difference in the performance of the unweighted and weighted ensembles (not shown here). We therefore restricted our analyses to an unweighted ensemble model.

We first drew 10000 samples from the posterior distribution of 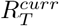 and forecasts from each model and then sampled each posterior distribution with equal weight to build the ensemble posterior distribution of 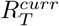 and corresponding forecasts.

### Short-term forecast horizon

The short-term forecast horizon was set to be 1 week. We produced forecasts for the week ahead (Monday to Sunday) using the latest data up to (and including) Sunday. We did not model the potential changes in the population immunity levels as any such change is not expected to affect the trajectory of the epidemic over this short time horizon.

### Medium-term forecasts

Over the course of the epidemic, the effect of the potential depletion of the susceptible population on the trajectory of the epidemic may become more pronounced. Inherently, by estimating transmissibility in real-time, the models outlined above account for any general decrease in the proportion of population being susceptible. However, the forecasts produced do not account for any further decrease in this proportion, which may become substantial when forecasting over a medium- to long-term time horizon.

### Estimating transmissibility for medium-term forecasts

To better capture ongoing trends in transmissibility over time, we examined changes in past ensemble estimates of 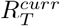 in consecutive weeks. To forecast over a time horizon longer than a week, we developed a novel heuristic that uses, in addition to the estimates of 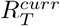, estimates from earlier weeks which are combined into a single weighted estimate 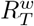 as follows. Starting with the estimates of 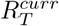, we went back one week at a time, for as long as the 95% credible interval (CrI) of 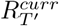 (where *T′ < T*) overlapped the 95% CrI of 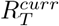. We then sampled from the posterior distribution of 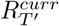 in each of those weeks, with a probability that decays exponentially in the past to favour the more recent estimates. That is, the distribution of 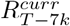 contributed to 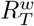 with a relative weight proportional to *e*^*−βk*^ (Fig. 1d). Each week, the rate of decay *β* was optimised by minimising the relative error in the predictions for the previous week.

### Accounting for depletion of the susceptible population due to naturally-acquired immunity

As the weighted reproduction number 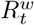 already accounts for the population immunity at time *t*, we first estimated an effective reproduction number 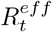, defined as the reproduction number if the entire population were susceptible. That is,

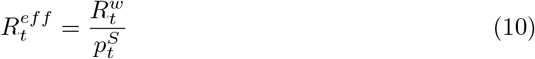

where 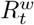 is the weighted reproduction number at time *t* and 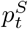 is the proportion of population that is susceptible to infection at time *t*. 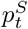 is given by 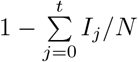 where *I*_*j*_ is the number of infections at time *j* and *N* is the total population. In estimating the potential future population immunity using this formulation, we only accounted for naturally acquired immunity assuming that the immunity acquired after infection persists. Since we were forecasting deaths (rather than infections), the true number of infections was estimated using a country-specific age-distribution weighted Infection Fatality Ratio (IFR) (SI Sec. 3).

We then incorporated the effect of a declining proportion of susceptible population due to naturally acquired immunity as

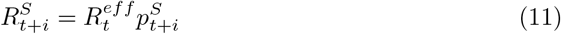

From the ensemble estimates of 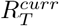, we first estimated 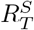. The medium-term forecasts were then produced using the renewal equation (Eq. (1)) and the forecasts used to update the estimates of 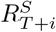 for each *i* ≥ 1 over the forecast horizon.

### Medium-term forecast horizon

The medium-term forecasts were made over a 4-week horizon using 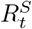. Since estimates of the weighted reproduction number could only be obtained once we had sufficient weekly estimates to combine, medium-term forecasts were produced from 29^th^ March to 29^th^ November 2020.

## Epidemic Phase

Following Abbott et al. [41], we defined the epidemic phase in a country at time *t* using the distribution of the reproduction number at *t* (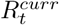 for the short-term forecasts and 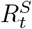 for the medium-term forecasts). At time *t*, we defined the epidemic phase in a country to be:

- ‘definitely growing’ if 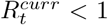 in less than 5% of the samples of the posterior distribution;
- ‘likely growing’ if 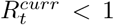 in less than 20% of the samples of the posterior distribution;
- ‘definitely decreasing’ if 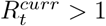 in less than 5% of the samples of the posterior distribution;
- ‘likely decreasing’ if 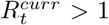 in less than 20% of the samples of the posterior distribution;
- ‘indeterminate’ otherwise.

### Assessing model performance

The model forecasts were validated against observed deaths as these became available. To quantitatively assess the performance of the model for both short- and medium-term forecasts, we used the mean relative error (MRE) and the coverage probability i.e. the proportion of observations that are in a given credible interval of the distribution of forecasts (SI Sec. 5). Following Gu [42], we compared the absolute error of the model (the absolute difference between the forecasts and observations averaged across simulated trajectories) with the error of a model that used (i) the average of the last 10 observations as a forecast for the week ahead (no growth), and (ii) forecasted using a linear model fitted to the last 10 observations (linear growth).

## Data and epidemiological parameters

We used the number of COVID-19 cases and deaths reported by the World Health Organisation (WHO) [1]. Any data anomalies were corrected using data published by the European Centre for Disease Prevention and Control [43], or other sources (SI Sec. 2). For the weekly analysis, we defined a country as having evidence of active transmission if at least 100 deaths had been reported, and at least ten deaths were observed in each of the past two weeks (SI Sec. 2). Countries with large variability in the reported deaths within each week over the analysis period were excluded from the final analysis for this work (SI Sec. 2 lists the full exclusion criterion). Results for 81 countries were included in the work presented here.

We assumed a gamma distributed serial interval with mean 6.48 days and standard deviation of 3.83 days following [44]. For simplicity, we assumed that the delay in reporting a death is the same as the delay from onset to a case being reported. We assumed that the delay in reporting of deaths follows a gamma distribution with mean of 10 days, and standard deviation of 2 days. These figures are roughly consistent with an onset-to-death delay of 15.9 days [45] and onset-to-diagnosis delay of 6.6–6.8 days [46]. The serial interval and delay distributions were discretised using R package EpiEstim [33]. We used a country-specific population-adjusted IFR estimated using the IFR reported in the REACT study (SI Sec. 3).

## Results

### Short-term forecasts and model performance

Beginning 8^th^ March 2020, we produced weekly forecasts for every country with evidence of sustained transmission. As the pandemic rapidly spread across the world, the number of countries included in the weekly analysis grew from 3 in the first week (week beginning 8^th^ March 2020), to 94 in the last week of analysis included in this study (week beginning 29^th^ November 2020) (SI Fig. 1).

Overall, the ensemble model performed well in capturing the short-term trajectory of the epidemic in each country. Across all weeks of forecast and all countries, an average 58.7% (SD 32.4%) of the observations were in the 50% credible interval (CrI) and 89.4% (SD 21.7%) of the observations were in 95% CrI (for a breakdown by country and week of forecast see SI Sec. 5.5).

The MRE across all countries and all weeks was 0.4 (SD 0.4) (Fig. 3). That is, on average the model forecasts were 0.4 times lower or higher than the observed incidence. In most countries, the reporting of both cases and deaths through the week was variable, with fewer numbers reported on some days of the week (typically, Saturday and Sunday). The variability in reported deaths strongly influenced the model performance. The MRE scaled linearly with the coefficient of variation (ratio of the standard deviation to the mean) in the reported deaths for the week of forecasting. Thus, the error in forecasts was on average similar to the variability in the reported deaths (SI Fig. 6). The MRE of the model scaled inversely with the weekly incidence i.e. the error was relatively large when the incidence was low (SI Fig. 6), as estimates of reproduction number when the incidence is low are inherently more unstable [47].

**Fig 2.**
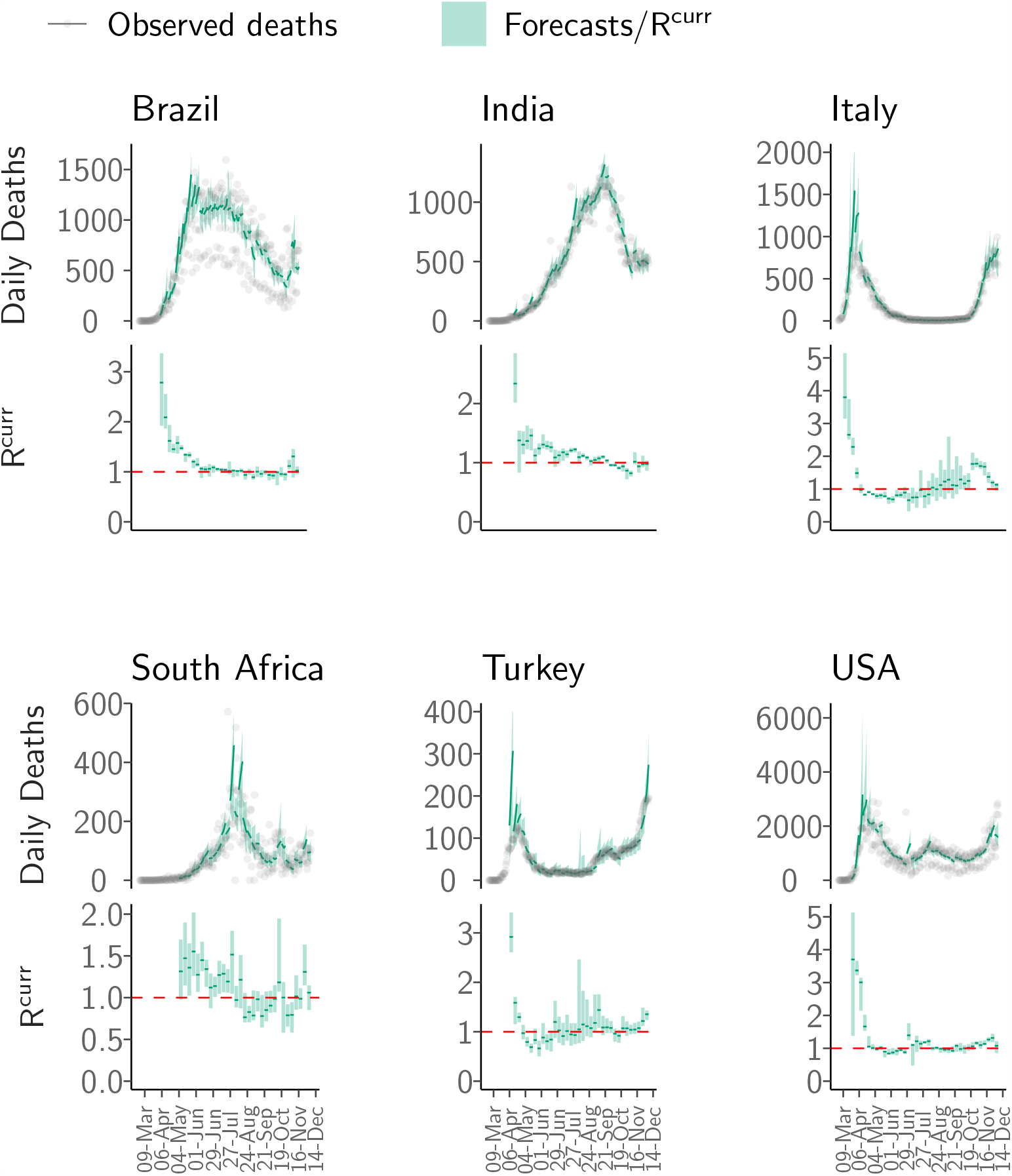
Short-term forecasts. The short-term forecasts and observed deaths for six countries: Brazil, India, Italy, South Africa, Turkey and the United States of America (USA). For each country, the top panel shows the observed deaths in gray; the solid green line shows the median forecast. The shaded interval represents the 95% CrI of forecasts. The forecasts were produced using the most recent estimates of 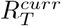 assuming that the transmissibility remains constant. The bottom panel for each country shows the effective reproduction number 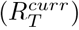 used to produce the forecasts. The solid green line in the bottom panel for each country is the median estimate of 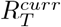 while the shaded region represents the 95% CrI. The dashed red line indicates the 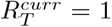 threshold. Note that the y-axis is different for each subfigure. See SI 2 for results for all other countries.

**Fig 3.**
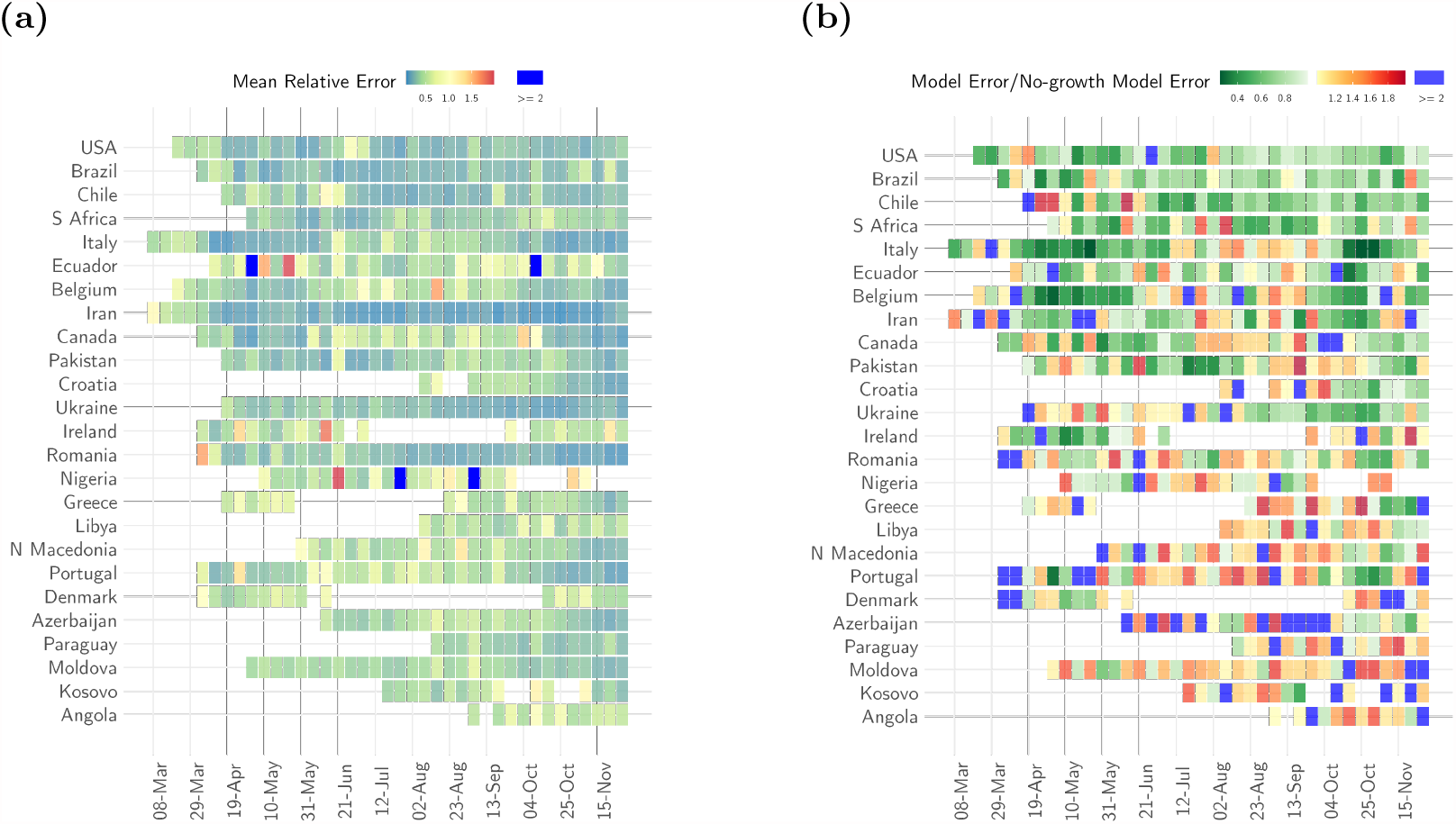
Short-term forecasts MRE and comparison with null model. (a) The mean relative error of the ensemble model for each week of forecast (x-axis) and for each country (y-axis). Dark blue cells indicate weeks where the relative error of the model was greater than 2. (b) The ratio of the absolute error of the model to the absolute error of a no-growth null model that uses the average of the last 10 days as a forecast for the week ahead. Shades of green show weeks for a given country where the ratio was smaller than 1 i.e., the ensemble model error was smaller, and weeks where the ratio was greater than 1 i.e. the ensemble model error was larger than the null model error are shown in shades of red (yellow to red). Dark blue indicates weeks when the ratio was larger than 2. In order to present a representative sample, we first ranked all countries by the percentage of weeks in which ensemble model error was smaller than the null model error. We then selected every third country from the top 75 countries in this list. Results for the selected 25 countries are shown here. See SI Fig. 4 for the results for other countries. Ordering of countries in the figure reflects the order in the ranked list i.e. countries with the highest percentage of weeks with model error smaller than null model error are shown on the top.

The model performance was largely consistent across epidemic phases (growing, likely growing, decreasing, likely decreasing and indeterminate) with similar coverage probability and MRE (SI Table 1). The slightly larger proportion of observations in the 50% and 95% credible intervals for the ‘indeterminate’ phase and the larger MRE in this phase together suggest that the model was ‘under-confident’ with large credible intervals [48].

We compared the performance of the model with that of a null no-growth model. In most instances, the ensemble model outperformed the null model. In 80.9% of the weeks in ‘definitely decreasing’ phase and 61.4% of weeks in ‘definitely growing’ phase, the absolute error of the model was smaller than the error made by the null model (Fig. 3, SI Sec. 5.2, SI Table 2). The null model performed better when the trajectory of the epidemic in a country was relatively stable exhibiting little to no change over the time frame of comparison. This is to be expected as the null model describes precisely this stable dynamic. Indeed, in 68.1% of the weeks in the ‘likely growing’ phase and 67.1% weeks classified as ‘indeterminate’ phase, the absolute error of the model was larger than the error made by the null model. However, the relative error of the model remained small even in countries and weeks where it did not perform as well as the null model. Similarly, our model performed better than a linear growth model across all phases, specifically in 96.4% of the weeks in ‘definitely decreasing’ phase and 70.3% weeks in ‘definitely growing’ phase (SI Sec. 5.3, SI Table 2).

### Medium-term forecasts and model performance

The rapidly changing situation and the various interventions deployed to stem the growth of the pandemic make forecasting at any but the shortest of time horizons extremely challenging [49]. Despite these challenges, we find that our medium-term forecasts were able to robustly capture the epidemic trajectory (Fig. 4) in all countries included in the analysis (4).

**Fig 4.**
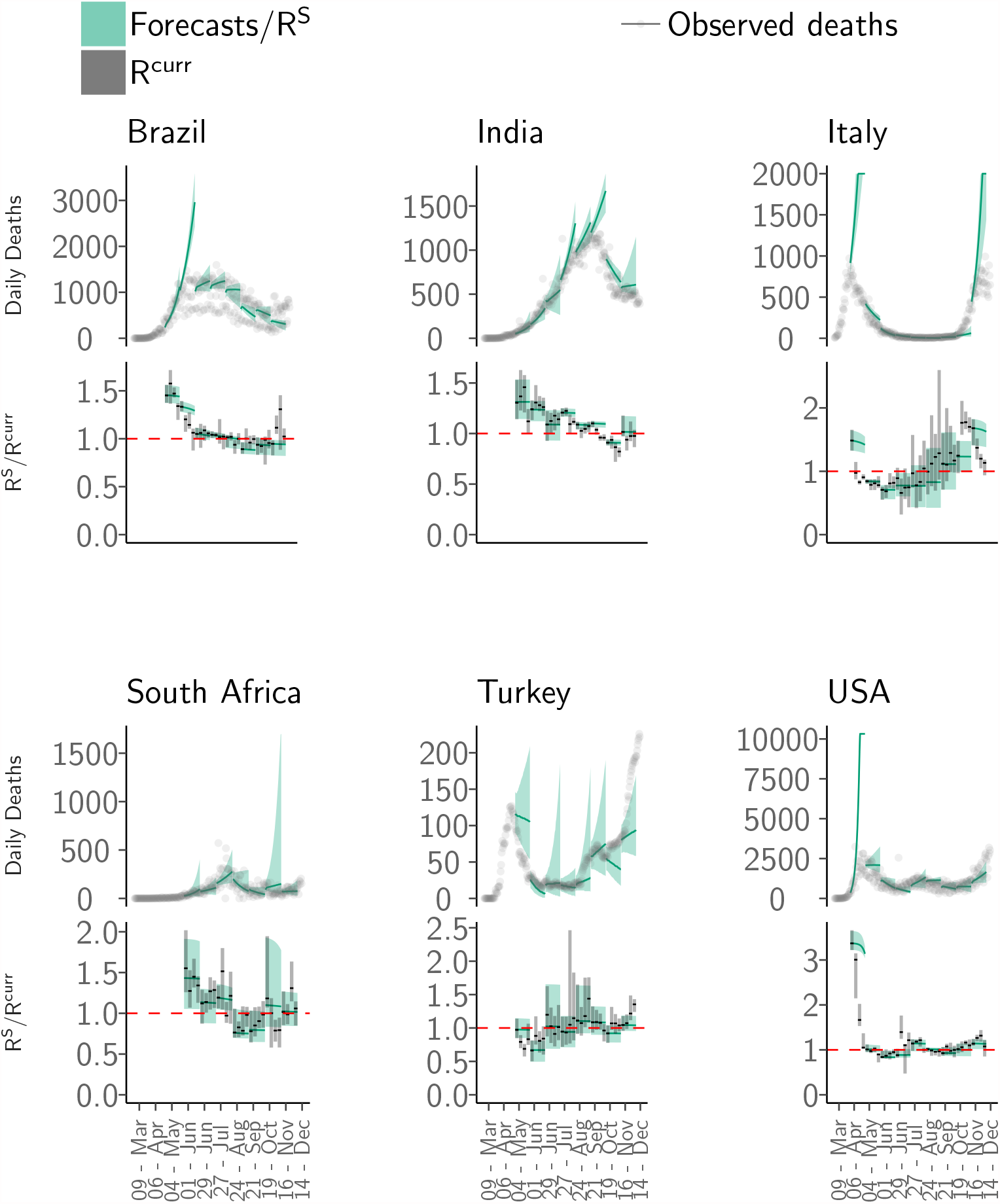
Medium-term forecasts. The medium-term forecasts and observed deaths for six countries: Brazil, India, Italy, South Africa, Turkey and the United States of America (USA). For each country, the top panel shows the observed deaths in grey; the solid green line shows the median the 4-weeks ahead forecast. The shaded interval represents the 95% CrI of forecasts. The bottom panel for each country shows the median (solid black line) and the 95% CrI (grey shaded area) of weekly estimate of 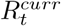 from the short-term forecasts and the median (green line) and the 95% CrI (shaded green area) of 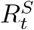 i.e. the reproduction number accounting for depletion of susceptible population from the medium-term forecasts over a 4-week horizon (Methods). The dashed red line indicates the 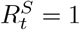 threshold. Note that the y-axis is different for each subfigure. The forecasts were produced every week over a 4-week forecast horizon. The figure shows all non-overlapping forecasts over the course of the pandemic. See SI 2 for results for all other countries and weeks.

Overall, the MRE remained small over a 4-week forecast horizon, with errors increasing over the projection horizon (SI Sec. 6.1). We therefore restricted the projection horizon to 4 weeks. The MRE across all countries in 1-week ahead forecasts was 0.4 (SD 0.3), increasing to 2.6 (SD 28.3) in 4-week ahead forecasts (Fig. 5, SI Fig. 10). The MRE for 1-week ahead forecasts was less than 1 (indicating that the magnitude of the error was smaller than the observation) in 91.1% of weeks for which we produced forecasts. The corresponding figure for 4-week ahead forecasts was 66.0% (SI Table 3).

**Fig 5.**
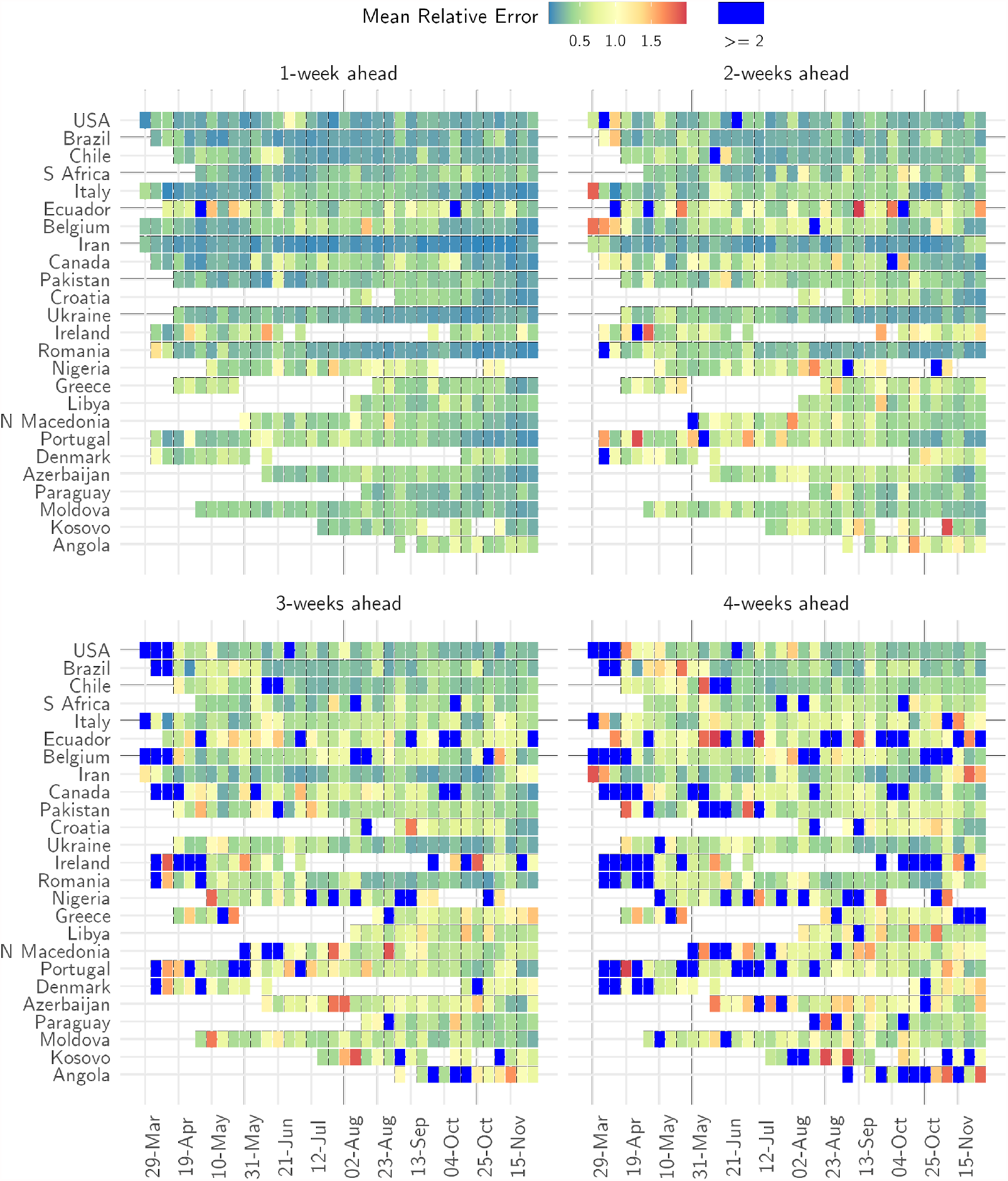
Relative error of medium-term forecasts. The mean relative error of the model in 1-week, 2-week, 3-week and 4-week ahead forecasts for each week when a forecast was made (x-axis) and for each country (y-axis). Blue cells indicate weeks where the relative error of the model was greater than 2. For ease of presentation, results are shown for the same 25 countries as Fig. 2. See SI Sec. 5 for the results for other countries.

The proportion of observations in the 50% CrI remained consistent across the forecast horizon and varied from 56.3% (SD 33.4%) in 1-week ahead forecasts to 45.6% (SD 40.9%) in 4-week ahead forecasts (SI Fig. 11, SI Fig. 12).

More importantly, using 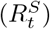 estimates from Eq. (11), we accurately characterised the phase of the epidemic in each country. Across the 81 countries and 2210 weeks (15470 days) for which we produced both short- and medium-term forecasts, the phase definition using the reproduction number estimates from medium-term forecasts 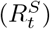 was consistent with that using the estimates from the short-term forecasts 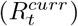 in 87.6% (13559/15470) of country-days (number of countries X number of days for which we produced forecasts. The phase definition using reproduction number estimates from medium-term forecasts was updated each day over the forecast horizon while the short-term forecasts assigned the same phase to all days of a week.) in 1-week ahead forecasts and in 84.9% (13138/15470) of country-days in 4-week ahead forecasts (Fig. 6). When the phase definitions using 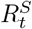 and 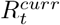 were different, the medium-term estimates most frequently misclassified them as a phase with greater uncertainty. For instance, in 253 weeks when the epidemic phase was identified as ‘definitely decreasing’ using weekly estimates and incorrectly characterised using medium-term estimates, it was misclassified as ‘likely decreasing’ in 100% (253/253 weeks) of country-days. Similarly, in the misclassified weeks, when the epidemic phase using weekly transmissibility estimates was ‘definitely growing’, the medium-term classification was ‘indeterminate’ in 43.7% (1175/2688) and ‘likely growing’ in 56.3% (1513/2688) of the country-days. This mischaracterisation is expected as the uncertainty in estimates of 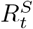 grows over the forecast horizon. Crucially, none of the weeks where 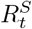 misclassified the epidemic phase, the phase using 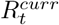 indicated the opposite trend (growing classified as decreasing or vice versa). This finding shows that the medium-term transmissibility estimates can be used a reliable indicator of the overall direction of the epidemic trajectory.

**Fig 6.**
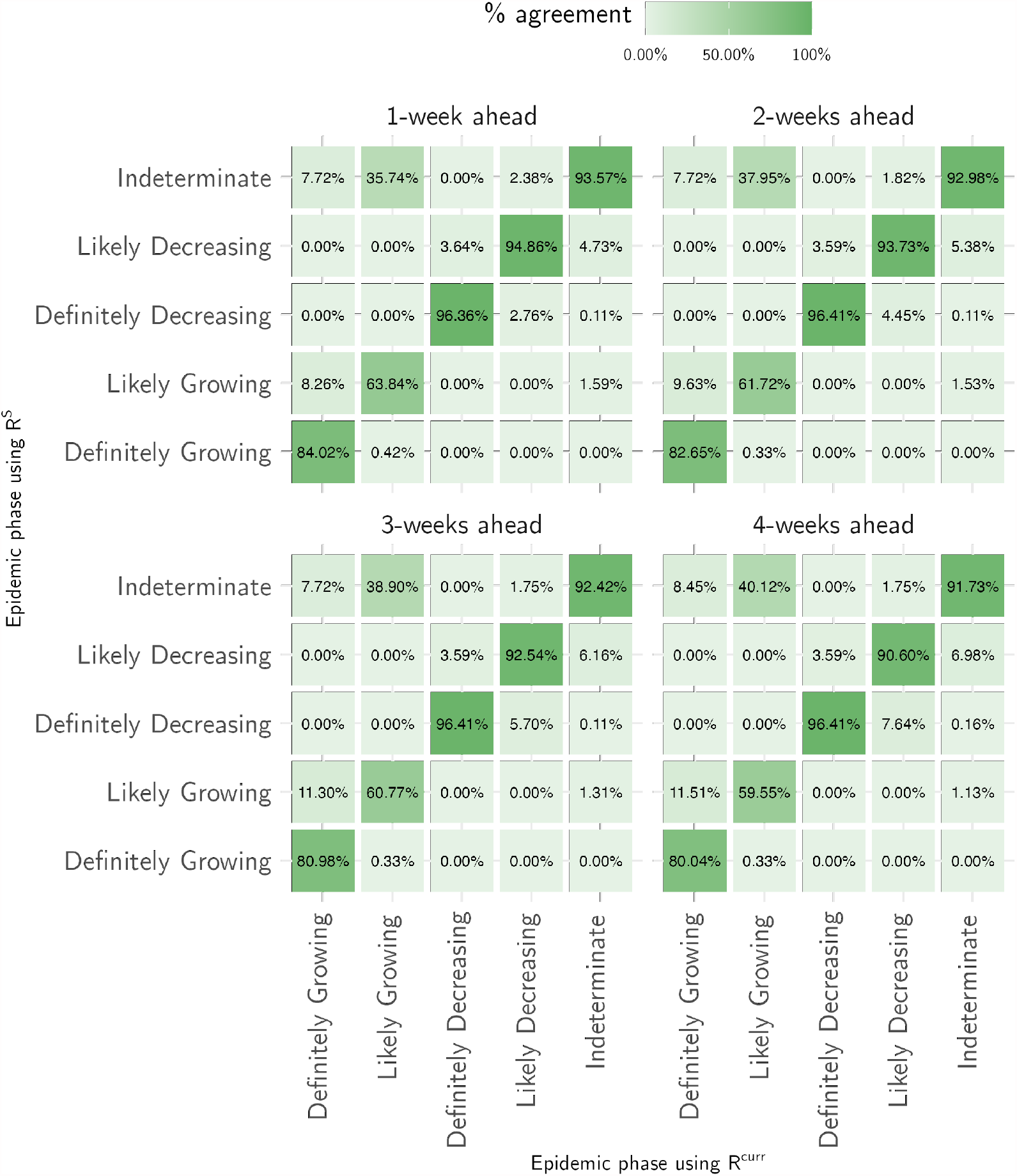
Characterisation of the epidemic phase. For a given classification of epidemic phase using the weekly estimates of the reproduction number from the short-term forecasts(x-axis), the figures in the cell show the percentage of days for which the characterisation was consistent using the medium-term reproduction number estimates (show on the y-axis)

## Discussion

Models used to forecast COVID-19 cases and/or deaths vary in complexity in the data used for model calibration. More complex and/or granular models rely on multiple data streams including data on hospital admissions and occupancy, testing, serological surveys and data on patient clinical progression and outcomes [21]. Such complex location-specific models can provide crucial insights into the ongoing epidemic and inform targeted public health interventions by synthesising evidence from different data streams. However, scaling such analysis to include multiple geographies is challenging because of the variability in availability and reliability of local surveillance data. The computational time needed to fit complex models make scaling them difficult and delays the timely provision of risk estimates.

In addition to the variable availability of surveillance data across countries, the wide-scale societal and behavioural changes brought about by the pandemic impose practical constraints on utilising data that are available for multiple countries. For instance, widely available data on the changes in mobility inferred from mobile phone usage released by Google and Apple were informative of the changes in transmission in the early phase of the COVID-19 pandemic and were used in several modelling studies [50, 51]. Although these data continue to be available, recent evidence suggests a decoupling of transmission and mobility in most countries [35, 52]. Models that relied on such additional data [51] or assumptions about non-pharmaceutical interventions [53] could not fit the observed trajectory well as the situation continued to change over the course of the epidemic.

Efforts to model and forecast COVID-19 transmission dynamics must therefore meet the challenges of a long and ongoing pandemic spread over an unprecedented scale. Modelling groups around the world have attempted to meet one or both challenges with various analyses conducted at a sub-national scale [54], at a national scale for a specific country [22, 55–57], and for several countries across the globe [41, 58, 59]. In contrast to models built for a region or country and calibrated using local data, models that aim to provide a global overview must be sufficiently general to describe the epidemic trajectory in a range of countries/regions using widely available data that are consistently available over time.

We have produced short-term forecasts and estimates of transmissibility for 81 countries for more than 65 weeks at the time of writing implementing three simple models that use only the time series of COVID-19 cases and deaths. We have thus traded particularity for generality, to allow us to carry out analysis for a large number of countries over a long period of time. As our methods make few assumptions and use routine surveillance data, they can be easily used during any other future outbreaks.

Despite the challenges inherent in forecasting a fast-moving pandemic in the presence of unprecedented public health interventions, our ensemble model was able to successfully capture the short-term transmission dynamics across all countries included in the analysis with small relative error in the weekly forecasts across different COVID-19 waves in each country. The variable performance of our model in weeks and countries with fewer deaths and/or large variability in reported deaths over weeks reflects this trade-off. In the absence of more detailed data, we assumed that epidemiological parameters such as the delay from onset of symptoms to death were the same across all countries and throughout the period of analysis. These parameters are likely to vary over time and between countries and using country-specific parameters could lead to moderate improvements in the model fits and forecast performance.

Due to the variability in testing and reporting of cases across different countries and over time within countries, using the reported number of cases to estimate transmissibility and produce forecasts is difficult without using more complex models. For these reasons, we primarily used deaths to estimate the reproduction number as we assumed that reporting of COVID-19 deaths was more complete and consistent over time and across different country surveillance systems. Although this assumption is unlikely to hold for many countries [60–62], our methods are robust to a constant rate of under-reporting over time as this would not alter the overall epidemic trends. A limitation of our work is that our estimates reflect the epidemiological situation with a delay of approximately 19 days (the delay from an infection to a death [53]). Nevertheless, our short-term forecasts and transmissibility estimates provide a valuable global overview and continuous insights into the dynamic trajectory of the epidemic in different countries. They also provide indirect evidence about the effectiveness of public health measures. Future research could investigate integrating more data streams into the models. In addition to the weekly reports that we publish, our work has also contributed to other international forecasting efforts [22, 48, 55].

We developed a simple heuristic to combine past estimates of transmissibility and a decline in the proportion of susceptible population to produce medium-term forecasts. We were able to achieve good model performance in forecasting up to 4 weeks ahead. Consistent with findings from other modelling studies [22], we found that the model error was unacceptably high beyond 4 weeks, suggesting that forecasting beyond this horizon is difficult. Importantly, our characterisation of the epidemic phase using weighted estimates of transmissibility were largely in agreement with that using short-term transmissibility estimates. Thus, our method was successful at capturing the broad trends in transmission up to 4 weeks ahead. The medium-term forecasts can therefore serve as a useful planning tool as governments around the world plan further implementation or relaxation of non-pharmaceutical interventions.

Our method incorporates the depletion of susceptible population and hence can in principle be extended to account for increasing population immunity as vaccination is rolled out across the world. However, inclusion of vaccine induced immunity depends on the availability of reliable data on vaccination coverage. Further, even if such data were available, teasing apart the impact of vaccination on transmission and mortality could be non-trivial. In light of these issues, it might be challenging to extend our approach to rigorously assess the effect of vaccination on epidemic trajectory on a global scale. However, our latest estimates of transmissibility indirectly reflect the impact of vaccination on transmission, allowing for the delay from vaccination to full immunity, and from infection to death. As we continue to track COVID-19 transmissibility globally, any temporal changes in transmissibility would implicitly account for the changes due to differential vaccination coverage.

Mathematical modelling and forecasting efforts have supported data-driven decision making throughout this public health crisis. Our work has aimed to improve global situational awareness. Using relatively simple approaches, we were able to produce robust forecasts for COVID-19 in 81 countries and provide crucial and actionable insights. This effort is being continued [26] as the world continues to grapple with renewed waves of COVID-19 cases.

## Data Availability

All data used in this analysis are available online https://github.com/mrc-ide/covid19-forecasts-orderly

https://shiny.dide.imperial.ac.uk/covid19-forecasts-shiny/

## Supporting information

**S1 File. Additional results**. The supplementary file contains additional results on model performance.

**S1 2. Web tool**. An interactive web-tool available at https://shiny.dide.imperial.ac.uk/covid19-forecasts-shiny/ presents both short- and medium-term forecasts, and reproduction number estimates for all countries included in the analysis.

## Acknowledgements

The authors acknowledge funding from the MRC Centre for Global Infectious Disease Analysis (reference MR/R015600/1), jointly funded by the UK Medical Research Council (MRC) and the UK Foreign, Commonwealth & Development Office (FCDO), under the MRC/FCDO Concordat agreement and is also part of the EDCTP2 programme supported by the European Union. JW acknowledges research funding from the Wellcome Trust (grant 102169/Z/13/Z). SB acknowledges funding from the Wellcome Trust (219415). This study is partially funded by the National Institute for Health Research (NIHR) Health Protection Research Unit in Modelling and Health Economics, a partnership between Public Health England, Imperial College London and LSHTM (grant code NIHR200908); and acknowledges funding from the MRC Centre for Global Infectious Disease Analysis (reference MR/R015600/1), jointly funded by the UK Medical Research Council (MRC) and the UK Foreign, Commonwealth & Development Office (FCDO), under the MRC/FCDO Concordat agreement and is also part of the EDCTP2 programme supported by the European Union. The views expressed are those of the author(s) and not necessarily those of the NIHR, Public Health England or the Department of Health and Social Care.

