## Supporting Information 1 for "Global predictions of short- to medium-term COVID-19 transmission trends : a retrospective assessment"

### Supplementary Information for Global predictions of short- to medium-term COVID-19 transmission trends: a retrospective assessment

July 19, 2021

#### Contents

**1 Overview** **2**

**2 Data** **2**

**3 Infection Fatality Ratio (IFR)** **4**

**4 Augmentation of observed cases for DeCa** **6**

**5 Model performance assessment** **8**

**7 Code** **27**

### 1 Overview

In this supplement, we present additional details on the data pre-processing (SI Sec. 2) and results on the performance assessment of the model for short- (SI Sec. 5) and medium-term forecasts (SI Sec. 6).

### 2 Data

#### Cleaning and pre-processing steps

We used the number of cases and deaths reported by the World Health Organisation (WHO) in the COVID-19 situation report [1]. If either the number of cases or deaths was negative for any country in WHO data, we used the corresponding figures from the data collated by the European Centre for Disease Prevention and Control [2] (if they were non-negative). If both these sources reported negative numbers, we replaced the negative count on a day with the average of the previous and subsequent 3 days. The deaths time series for each country was then visually inspected and any anomalies (e.g. when a large number of deaths were reported on a single day as a correction) were manually corrected using media reports or alternative sources. A complete list of corrections applied to the data is available on the github repository of this project (`https://github.com/mrc-ide/covid19-forecasts-orderly/blob/main/src/prepare_ecdc_data/prepare_ecdc_data.R`).

#### Inclusion/Exclusion Criteria

For the analysis carried out every week, we defined a country as having evidence of active transmission if at least 100 deaths had been reported in a country, and at least ten deaths were observed in the country in each of the past two weeks. Forecasts were produced every Monday for the week ahead (Monday to Sunday) using data reported up to the previous

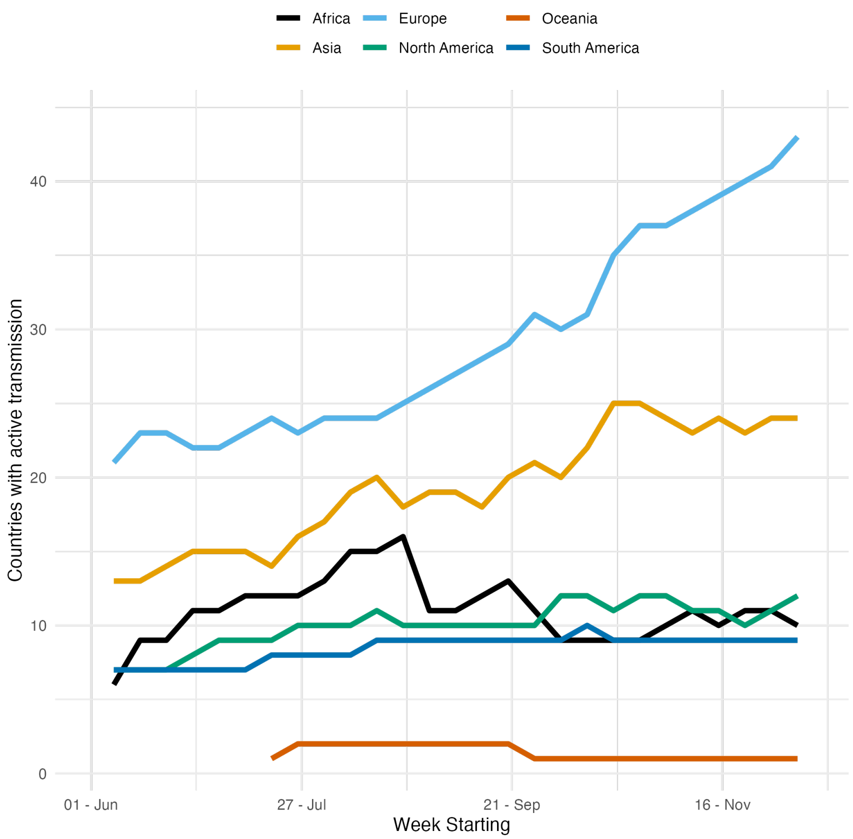

**Figure 1.** Number of countries included in the weekly reports from 8th March to 29th November 2020. the number of countries included in the weekly analysis grew from 2 in the first week (week starting 8th March 2020), to 94 in the last week of analysis included here (week starting 29th November 2020). Note that some countries that were included in the weekly reports have been excluded from the analysis presented in the manuscript if the average weekly coefficient of variation of the reported deaths between 8th March and 29th November 2020 was greater than 1.1.

day. Some countries were excluded from the analysis despite meeting these thresholds because the number of deaths per day did not allow reliable inference.

For the summary presented in this manuscript, we included all countries in the weekly analysis except countries with average weekly coefficient of variation (CV i.e. the ratio of standard deviation to the mean) of the reported deaths between 8th March and 29th November 2020 greater than 1.1 (the 60th quantile of the distribution of CV across all countries). This criterion resulted in the exclusion of 53 countries. 81 countries were included in the final analysis.

### 3 Infection Fatality Ratio (IFR)

To obtain a IFR distribution, we used the reported deaths and the estimated number of infections in age groups 15-44, 45-64, 65-74 years in the United Kingdom [3]. We first drew 10000 samples from a normal distribution with mean the estimated mean number of infections and standard deviation set to half the width of the 95% CI divided by 1.96. We divided the reported number of deaths in the corresponding age groups by the estimated number of infections to obtain age-disaggregated IFR distributions. We then obtained a country-specific IFR distribution as a weighted sum of the age-disaggregated IFR where the weights are the proportion of the total population in each group in a country [4].

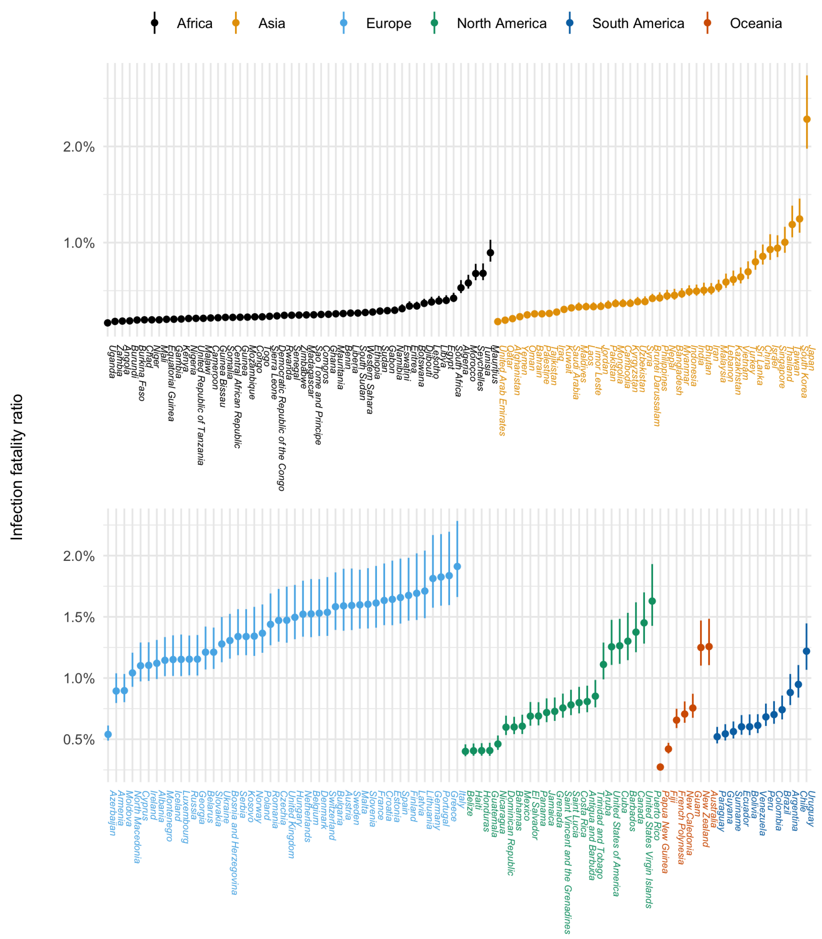

**Figure 2.** Population adjusted IFR distribution. The solid dots indicate the median estimate and the vertical bars represent the 95% CrI.

### 4 Augmentation of observed cases for DeCa

In the DeCa model, forecasts of deaths at time $t$ rely on the number of cases from the beginning of the time series to time $t$. We obtained a distribution of cases in the week for which we are producing forecasts by sampling from a gamma distribution with the mean and standard deviation equal to those of the most recent week of data on cases. We illustrate this process and also show that this does not influence the results under the chosen distribution of delays from case report to death (SI Fig. 3).

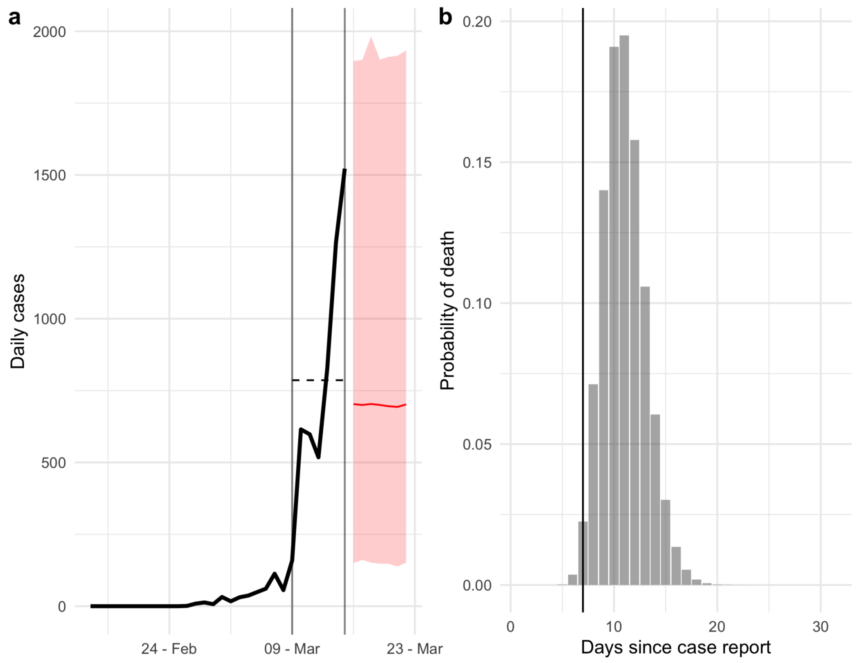

**Figure 3.** (a) The observed time series of cases (thick black line) is augmented by sampling from a gamma distribution with the mean and standard deviation of the cases in the most recent week of data. The vertical lines indicate the last week and the dashed horizontal line is the mean of the cases in this period. The red line and the shaded area represent the median and the 95% CrI of the sampled cases. (b) The probability distribution of delays from case report to death. For a case reported at time $t$, the probability of death within $d$ days is the sum of probabilities from 0 to $d$. In particular, the probability that a case will die within a week (our short-term forecast horizon) is the sum of probabilities to the left of the horizontal line (7 days), which is approximately 2%.

### 5 Model performance assessment

The following metrics were used to assess the model performance:

- **Mean relative error** The mean relative error (MRE) is a widely used measure of model accuracy [5]. The mean relative error for the forecasts $\hat{D}_t$ at time $t$ is defined as:

$$MRE_t(D_t, \hat{D}_t) = \frac{\sum_{s=1}^{N} |D_t - \hat{D}_{t,s}|}{N * (D_t + 1)},$$

  where $D_t$ denotes the observed deaths at time $t$, $N$ is the number of simulated trajectories and $\hat{D}_{t,s}$ denotes the $s^{th}$ simulation at time $t$ [6]. That is the mean relative error at time $t$ is averaged across all simulated trajectories and normalised by the observed incidence. We add 1 to the observed value to prevent division by 0. A MRE value of $k$ means that the average error is $k$ times the observed value.

- **Comparison with null model** Compare the absolute error made by the model with the absolute error made by a null model that uses the average of the last 10 observations as the forecast for the week ahead. We also compared the model error with the error made by a linear model (forecasts from a line fitted to the last 10 observations).

- **Coverage probability** Coverage probability refers to the proportion of observations that are contained in given credible interval (CrI) of the distribution of forecasts. For a well-calibrated model, 50% of the observations should be contained in the 50% CrI [7]. For a X% CrI, coverage probability higher than X% indicates that the model is under-confident while a value less than X% suggests that the model is over-confident with narrow CrIs.

For each country and for each week, the time series of observed deaths was first smoothed by taking a 3-day rolling mean. The average of the daily MRE was used as the weekly MRE.

#### 5.1 Mean relative error by epidemic phase

| Epidemic phase | Proportion in 50% CrI | Proportion in 95% CrI | MRE |
|---|---|---|---|
| Definitely decreasing | 48.5% (29.9%) | 83.1% (24.3%) | 0.4 (0.3) |
| Likely decreasing | 60.7% (33.1%) | 91.2% (20.2%) | 0.4 (0.3) |
| Definitely growing | 48.6% (31.9%) | 84.7% (24.7%) | 0.4 (0.7) |
| Likely growing | 62.2% (31.0%) | 92.4% (19.4%) | 0.5 (0.5) |
| Indeterminate | 66.1% (31.2%) | 92.4% (18.9%) | 0.5 (0.6) |

**Table 1.** Coverage probability and mean relative error of short-term forecasts in each epidemic phase. The values show the average of the metric across countries and weeks of forecast. The standard deviation is shown in parentheses.

#### 5.2 Relative error and comparison with no-growth model

This section presents the mean relative error of the model and comparison of the model error with the error made by a model that uses the average of the past 10 days as the forecast for the week ahead.

**(a)**

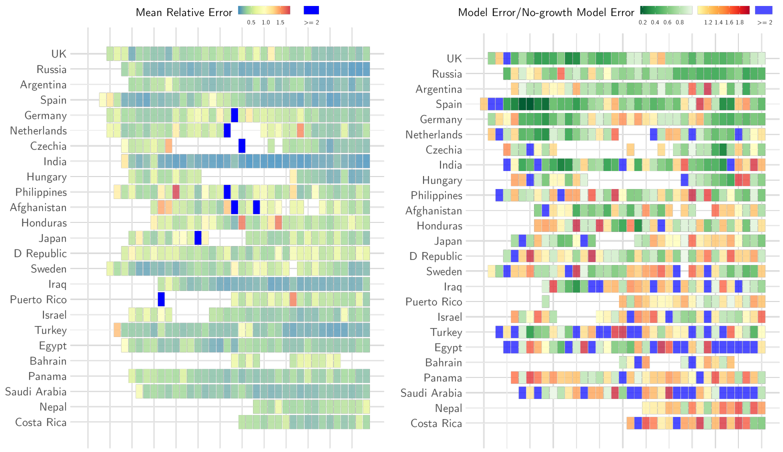

**(b)**

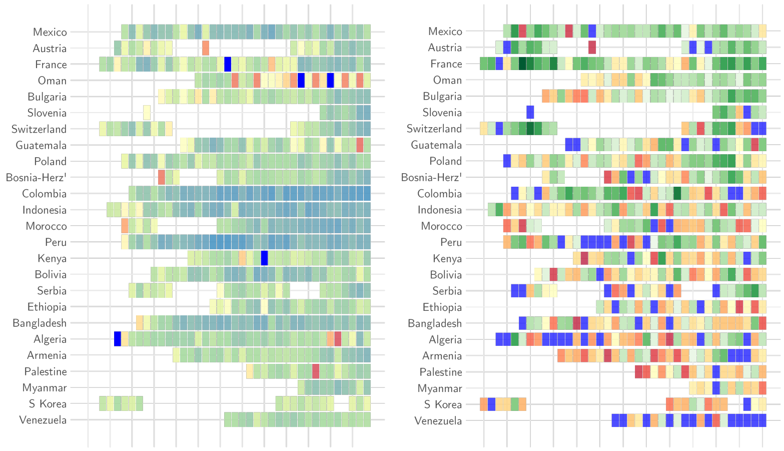

**(c)**

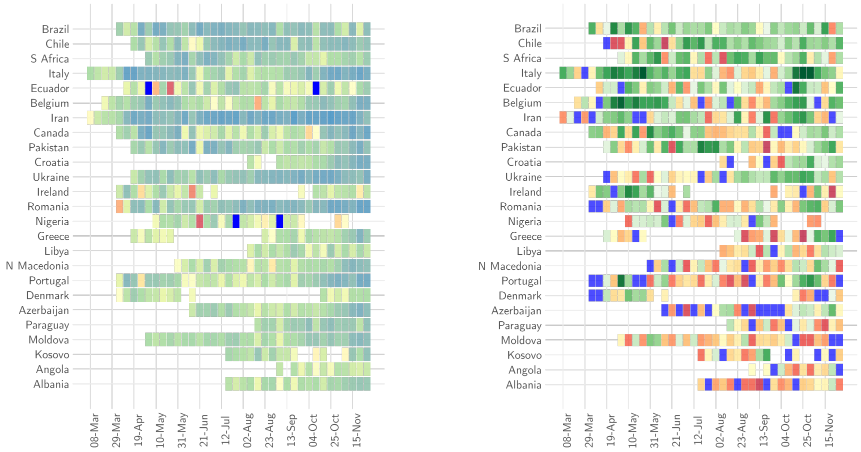

**Figure 4. Mean relative error and comparison with null model** In each panel, the left graph shows the relative error of the ensemble model for each week of forecast (x-axis) and for each country (y-axis). Dark blue cells indicate weeks where the relative error of the model was greater than 2. The right panel shows the ratio of the absolute error of the model to the absolute error of the no-growth null model. Shades of green show weeks for a given country where the ratio was smaller than 1 i.e., the model error was smaller, and weeks where the ratio was greater than 1 i.e. the model error was bigger than the null model error are shown in shades of red (yellow to red). Dark blue cells indicate weeks where the ratio was greater than 2. Panels (a) - (c) show results for all countries included in the analysis.

##### 5.2.1 Comparison with no-growth and linear models by phase

| Phase | Ensemble model error <No-growth model error | Ensemble model error <Linear model error | Weeks |
|---|---|---|---|
| Likely decreasing | 54.5% | 88.4% | 224 |
| Definitely decreasing | 80.9% | 96.4% | 251 |
| Likely growing | 31.9% | 74.8% | 301 |
| Definitely growing | 61.4% | 70.3% | 542 |
| Indeterminate | 32.9% | 80.7% | 887 |

**Table 2.** Comparison of the absolute error of the ensemble model with that made by a null no-growth model or a predictions from a linear model as forecast for the week ahead for each phase of the pandemic. The right-most column (Weeks) shows the total number of weeks in a given phase.

#### 5.3 Relative error and comparison with a linear model

This section presents the relative error of the ensemble model and comparison of the model error with the error of a linear model (a line fitted to the past 10 observations). The linear model was fitted in rstannarm [8] and the forecasts were sampled from the posterior predictive distribution.

**(a)**

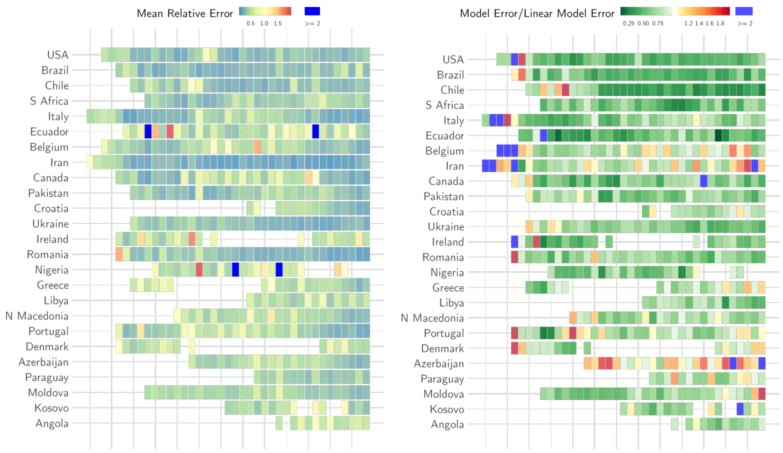
Mean Relative Error
0.5 1.0 1.5
>= 2
Model Error/Linear Model Error
0.25 0.50 0.75
1.2 1.4 1.6 1.8
>= 2
USA
Brazil
Chile
S Africa
Italy
Ecuador
Belgium
Iran
Canada
Pakistan
Croatia
Ukraine
Ireland
Romania
Nigeria
Greece
Libya
N Macedonia
Portugal
Denmark
Azerbaijan
Paraguay
Moldova
Kosovo
Angola

**(b)**

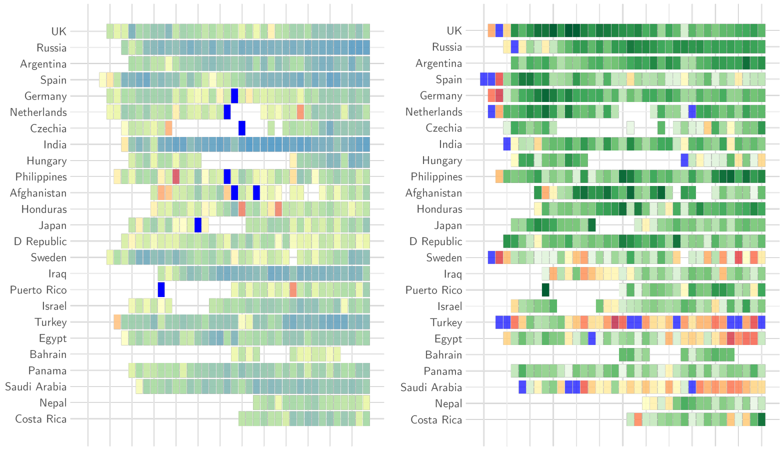
UK
Russia
Argentina
Spain
Germany
Netherlands
Czechia
India
Hungary
Philippines
Afghanistan
Honduras
Japan
D Republic
Sweden
Iraq
Puerto Rico
Israel
Turkey
Egypt
Bahrain
Panama
Saudi Arabia
Nepal
Costa Rica

**(c)**

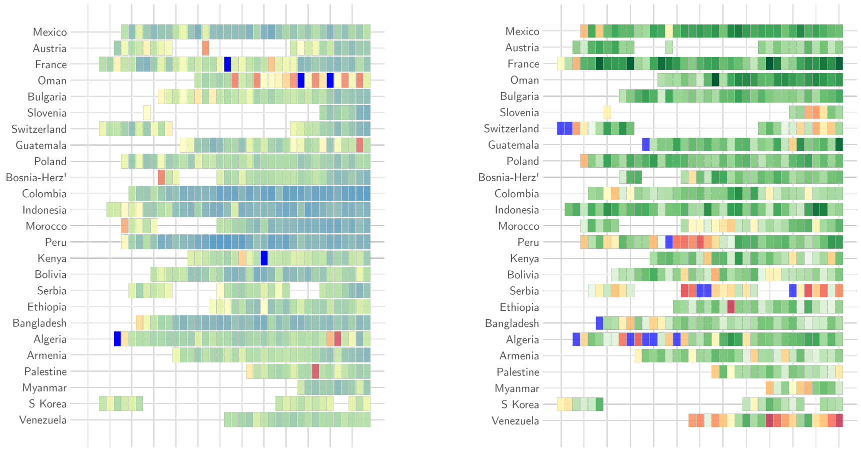

**(d)**

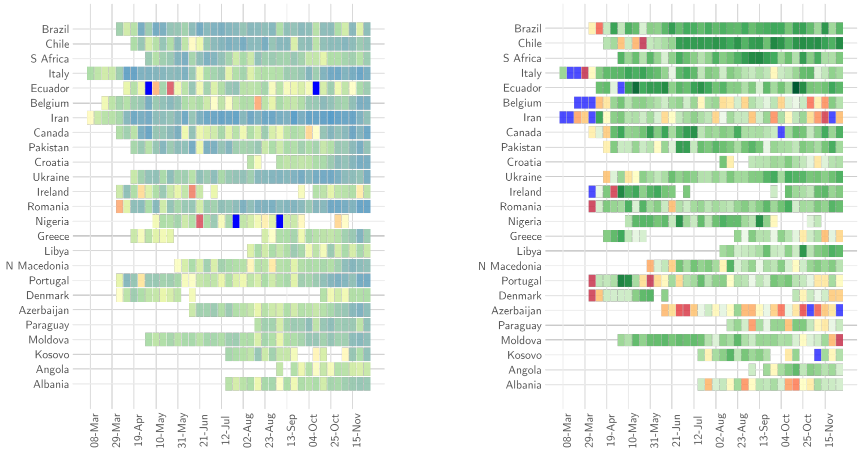

**Figure 5. Relative error and comparison with a linear model** In each panel, the left graph shows the mean relative error of the model for each week of forecast (x-axis) and for each country (y-axis). Dark blue cells indicate weeks where the relative error of the model was greater than 2. The right panel shows the ratio of the absolute error of the model to the absolute error of forecasts made using a linear model. Shades of green show weeks for a given country where the ratio was smaller than 1 i.e., the model error was smaller, and weeks where the ratio was greater than 1 i.e. the model error was bigger than the null model error are shown in shades of red (yellow to red). Dark blue cells indicate weeks where the ratio was bigger than 2. Panels (a)-(d) show results for all countries included in the analysis.

#### 5.4 Mean relative error compared with the weekly CV

The relative error of the model was proportional to the CV of the number of deaths reported each week and inversely proportional to the weekly incidence.

**(a)** **(b)**

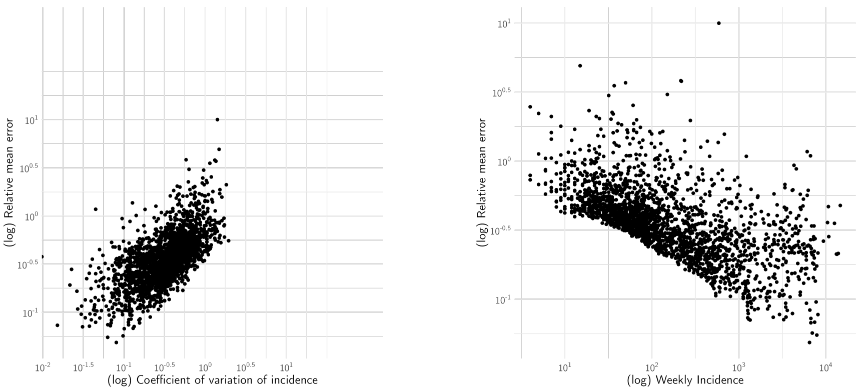

**Figure 6.** The log MRE scales linearly with the log weekly CV (a) and inversely with the log weekly incidence (b).

#### 5.5 Coverage Probability

This section presents the proportion of observations in 50% CrI and 95% CrI for each country and each week of forecast.

##### Proportion of observations in 50% CrI

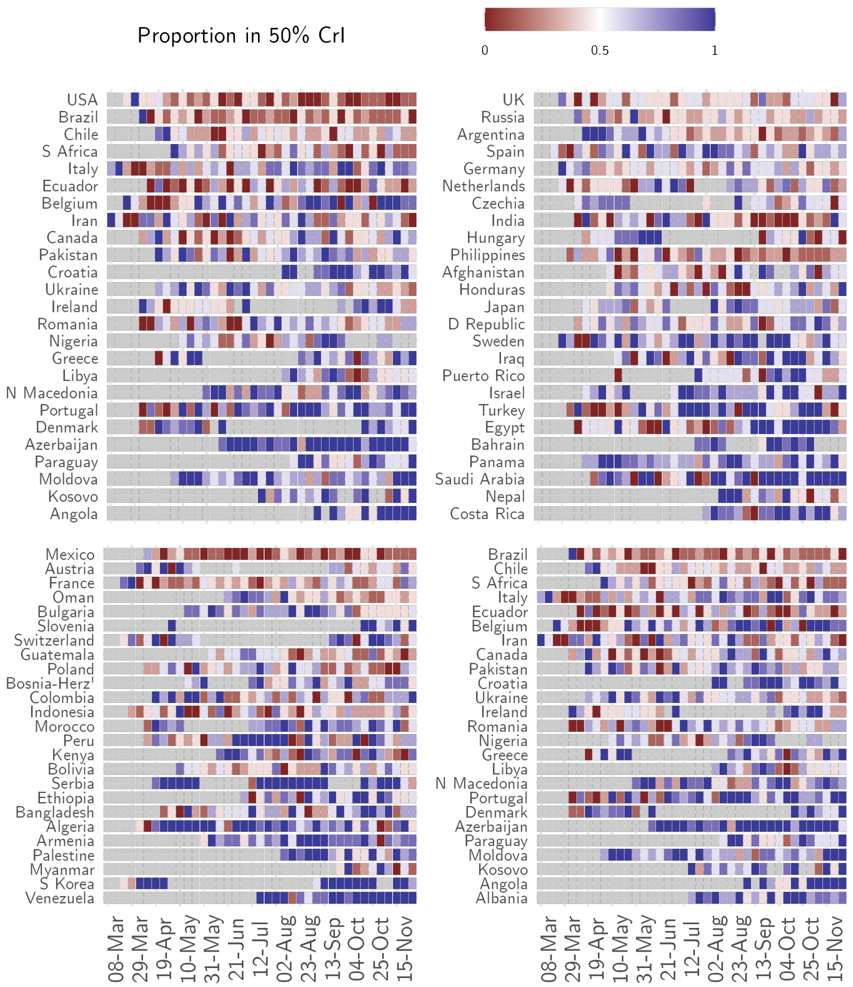

**Figure 7.** For each week of forecast (x-axis) and each country (y-axis), the proportion of observations in the 50% CrI of the forecasts. Gray cells indicate weeks where a country was not included in the analysis because the number of deaths did not meet the threshold (SI Sec. 2).

#### Proportion of observations in 95% CrI

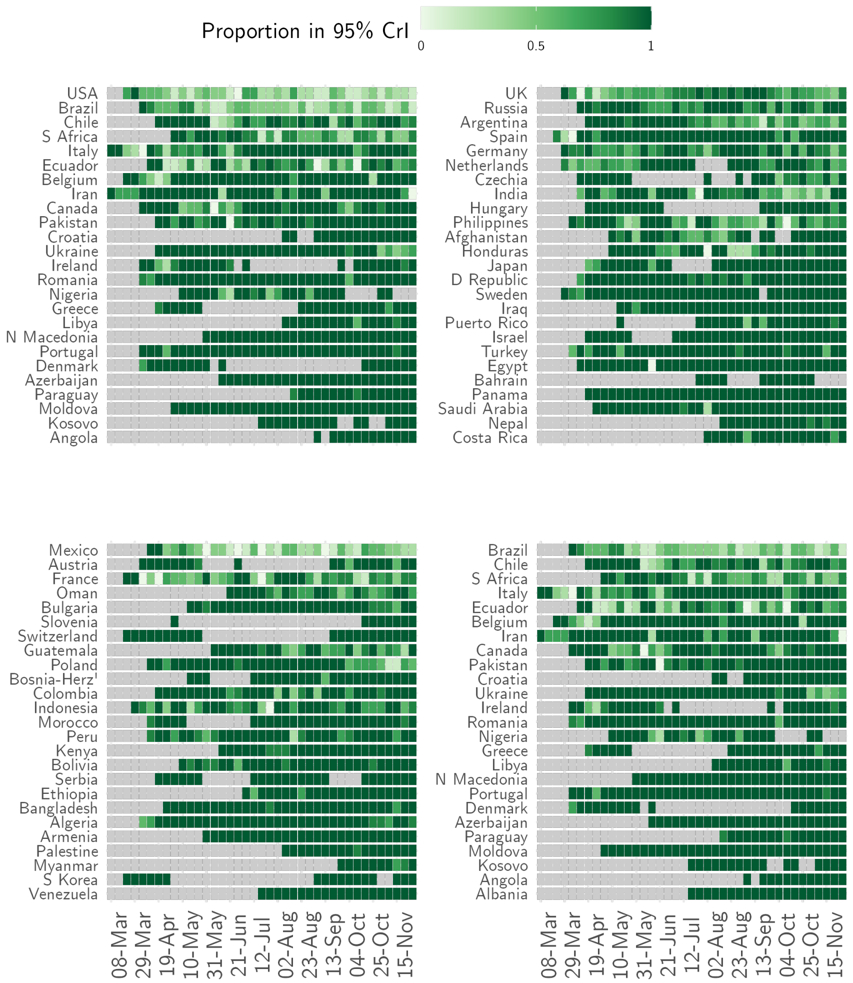

**Figure 8.** For each week of forecast (x-axis) and each country (y-axis), the proportion of observations in 95% CrI of the forecasts. Gray cells indicate weeks where a country was not included in the analysis because the number of deaths did not meet the threshold (SI Sec. 2).

### 6 Medium-term forecasts

This section presents the performance assessment results for medium-term forecasts. The relative error for each country and week of forecast are presented in (SI Sec. 6.1) and coverage probability are shown in (SI Sec. 6.2).

#### 6.1 Relative error

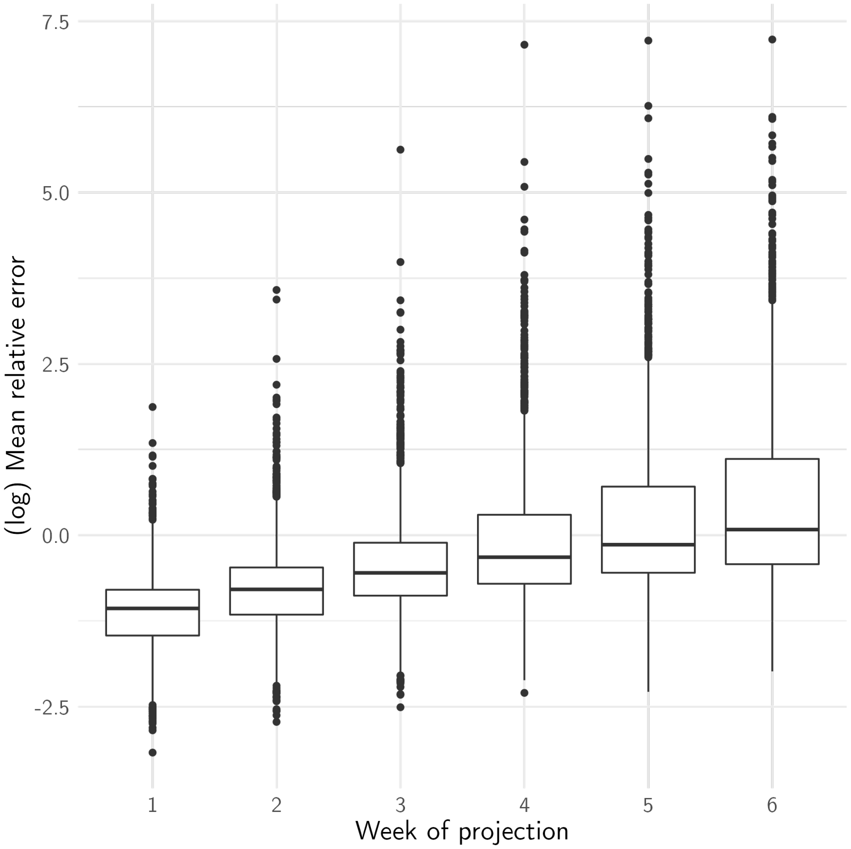

**Figure 9.** The mean relative error grew over the projection horizon becoming unaccpetably high beyond a 4-week horizon.

**(a)**

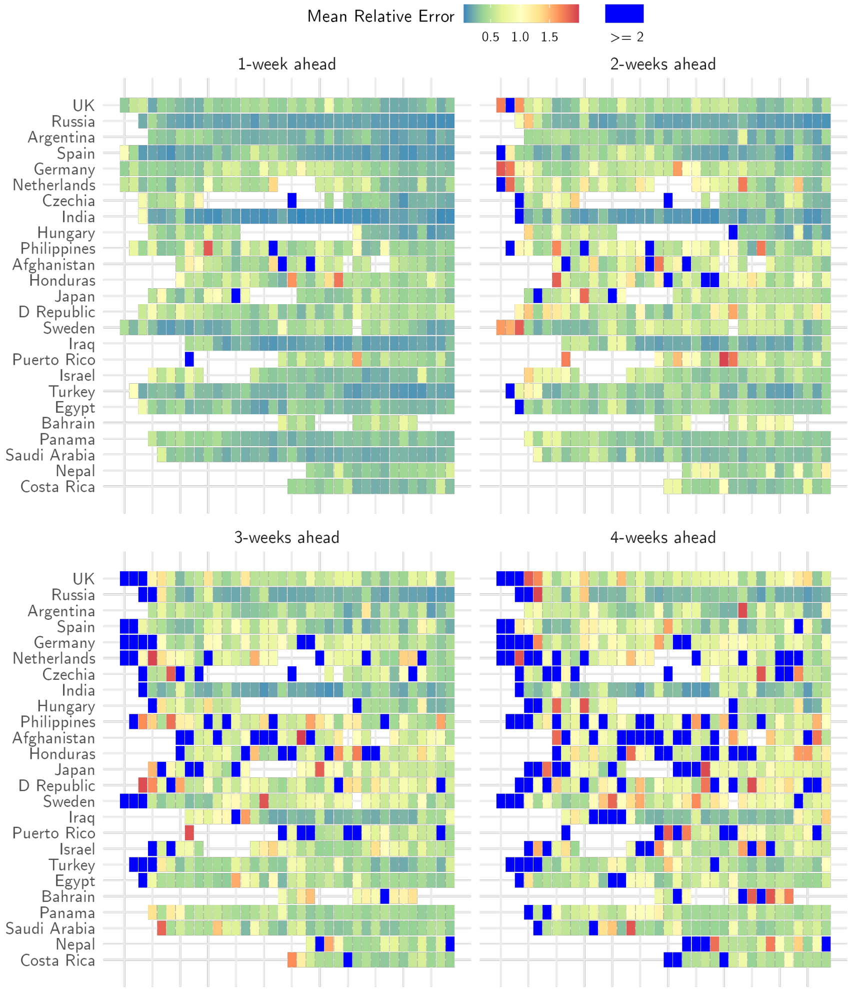

Mean Relative Error
0.5
1.0
1.5
>= 2
1-week ahead
2-weeks ahead
3-weeks ahead
4-weeks ahead
UK
Russia
Argentina
Spain
Germany
Netherlands
Czechia
India
Hungary
Philippines
Afghanistan
Honduras
Japan
D Republic
Sweden
Iraq
Puerto Rico
Israel
Turkey
Egypt
Bahrain
Panama
Saudi Arabia
Nepal
Costa Rica

**(b)**

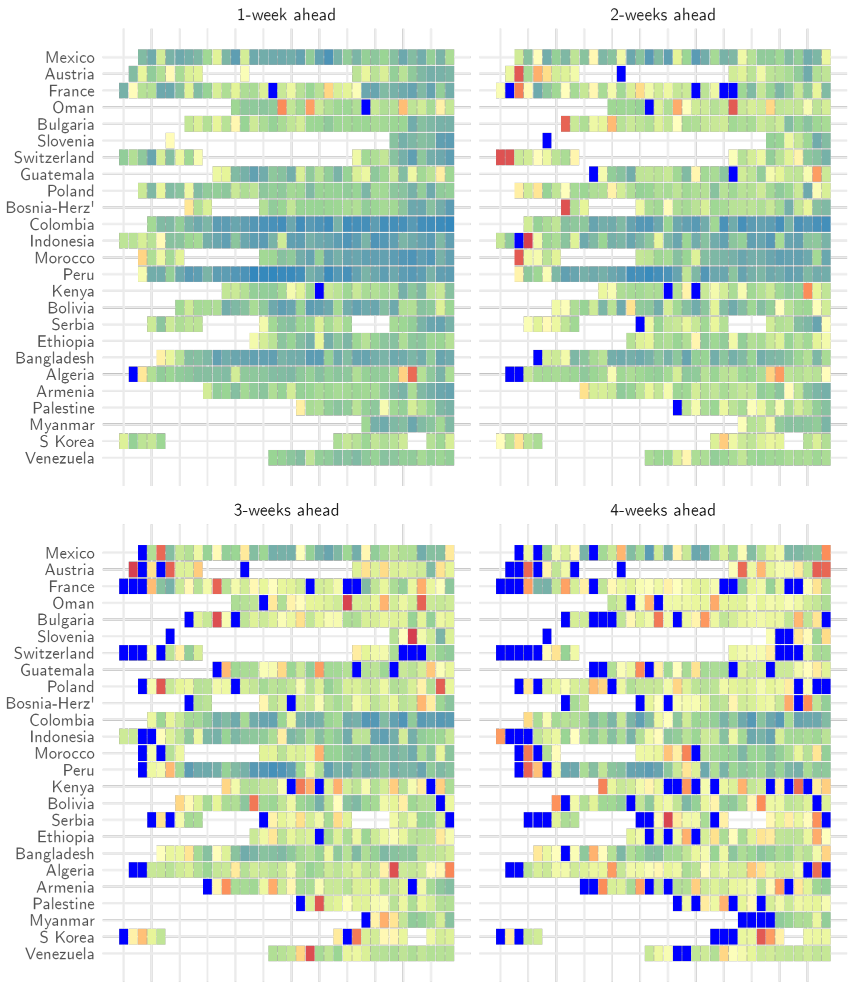
1-week ahead
2-weeks ahead
3-weeks ahead
4-weeks ahead
Mexico
Austria
France
Oman
Bulgaria
Slovenia
Switzerland
Guatemala
Poland
Bosnia-Herz'
Colombia
Indonesia
Morocco
Peru
Kenya
Bolivia
Serbia
Ethiopia
Bangladesh
Algeria
Armenia
Palestine
Myanmar
S Korea
Venezuela

**(c)**

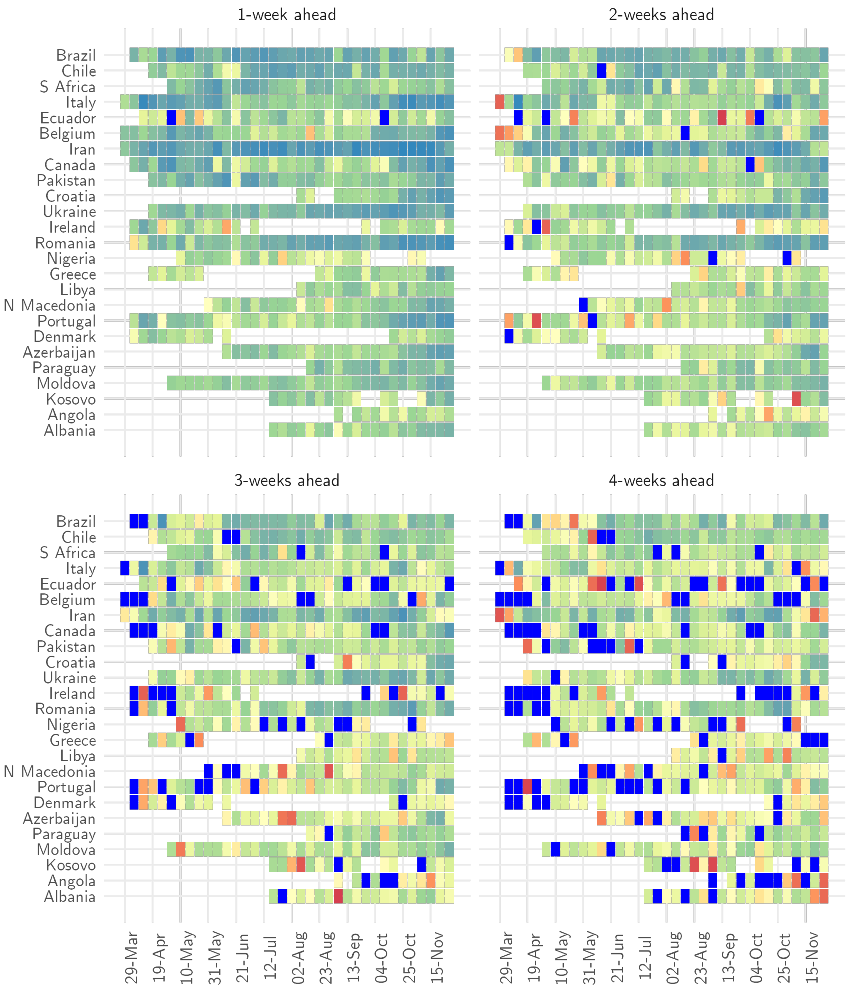

**Figure 10. Mean relative error of medium-term forecasts.** The relative error of the model in 1-week, 2-week , 3-week, and 4-week ahead forecasts for each week of forecast (x-axis) and for each country (y-axis). Dark blue cells indicate weeks where the relative error of the model was greater than 2. Panels (a)-(c) present results for all countries included in the analysis.

| Week of forecast | MRE <0.5 | MRE <1 |
|---|---|---|
| 1 | 80.8% | 91.1% |
| 2 | 58.3% | 89.5 % |
| 3 | 33.2% | 78.3% |
| 4 | 25.6% | 66.0% |

**Table 3. The MRE of medium-term forecasts remained relatively small over a 4-week forecast horizon. The MRE was less than 1 in 66.0% and less than 0.5 in 25.6% of weeks in 4-week ahead forecasts.**

#### 6.2 Coverage Probability

##### Proportion of observations in 50% CrI

**(a)**

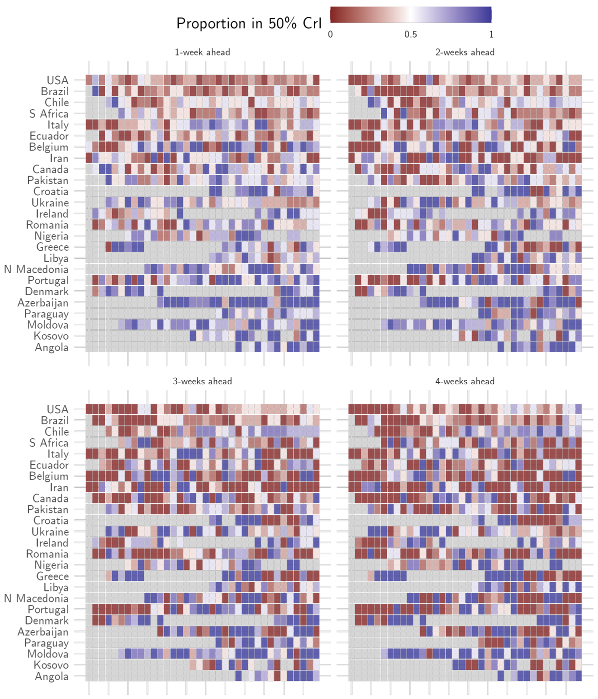
Proportion in 50% CrI
0
0.5
1
1-week ahead
2-weeks ahead
3-weeks ahead
4-weeks ahead
USA
Brazil
Chile
S Africa
Italy
Ecuador
Belgium
Iran
Canada
Pakistan
Croatia
Ukraine
Ireland
Romania
Nigeria
Greece
Libya
N Macedonia
Portugal
Denmark
Azerbaijan
Paraguay
Moldova
Kosovo
Angola

**(b)**

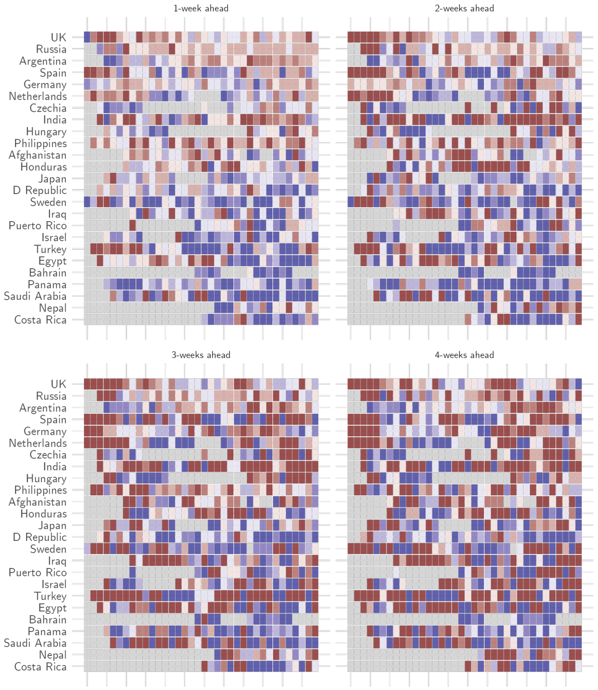
1-week ahead
2-weeks ahead
3-weeks ahead
4-weeks ahead
UK
Russia
Argentina
Spain
Germany
Netherlands
Czechia
India
Hungary
Philippines
Afghanistan
Honduras
Japan
D Republic
Sweden
Iraq
Puerto Rico
Israel
Turkey
Egypt
Bahrain
Panama
Saudi Arabia
Nepal
Costa Rica

**(c)**

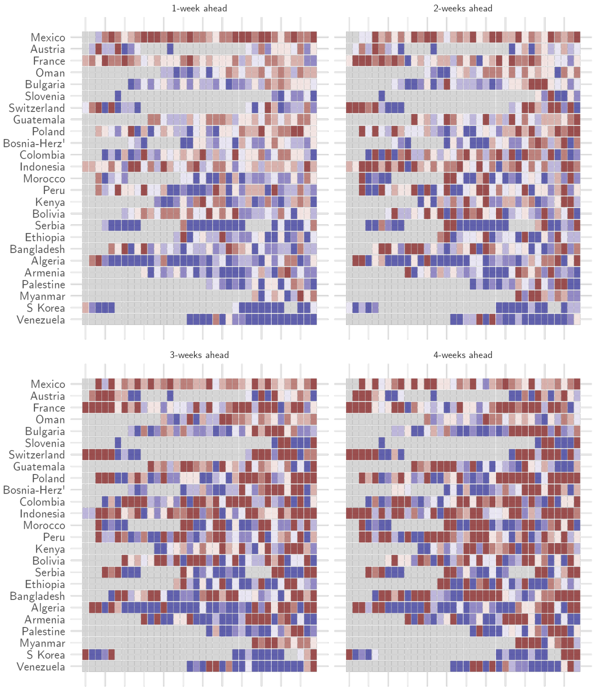

**(d)**

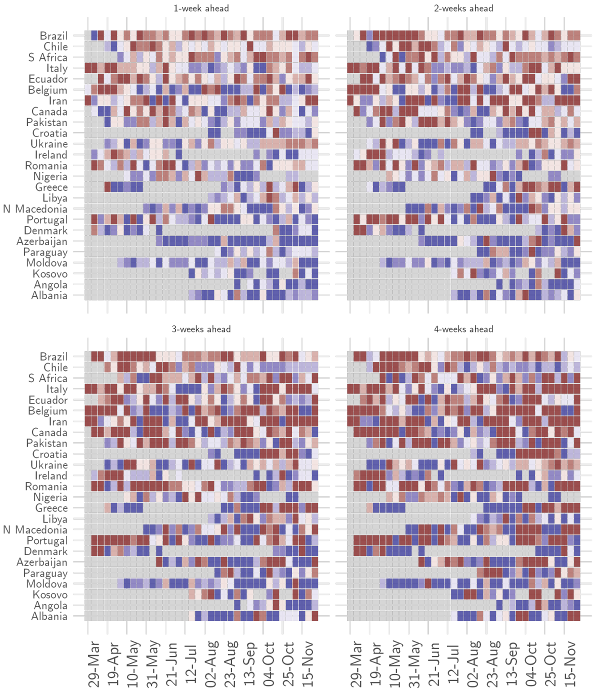

**Figure 11.** The proportion of observations in the 50% CrI of the forecasts for 1-week, 2-week , 3-week, and 4-week (clockwise from top left) ahead for each week of forecast (x-axis) and for each country (y-axis). Panels (a)-(d) present results for all countries included in the analysis. Gray cells indicate weeks where a country was not included in the analysis because the number of deaths did not meet the threshold (SI Sec. 2).

### Proportion of observations in 95% CrI

**(a)**

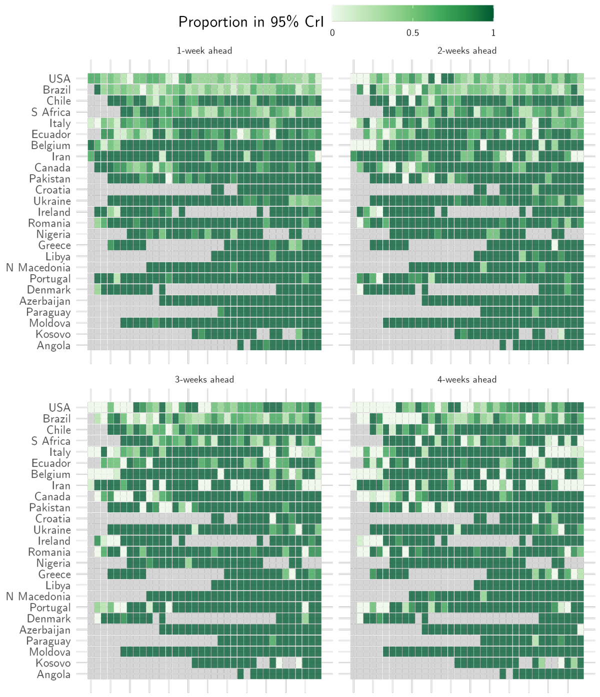

(b)

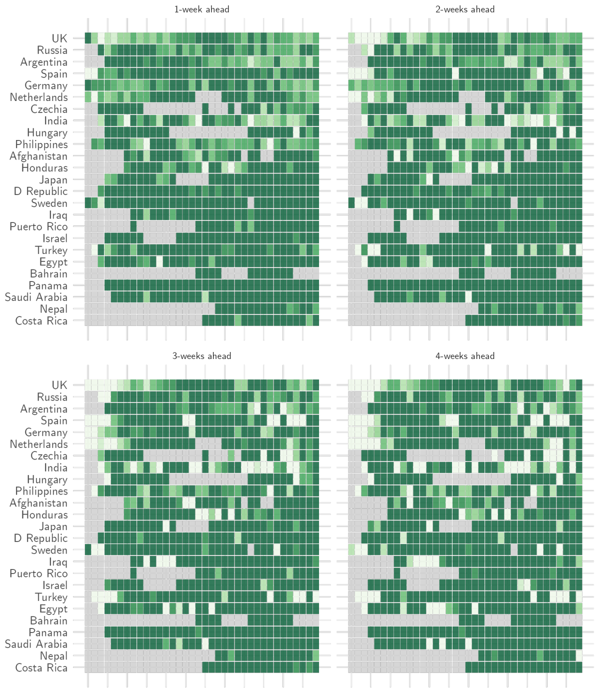
1-week ahead
2-weeks ahead
3-weeks ahead
4-weeks ahead
UK
Russia
Argentina
Spain
Germany
Netherlands
Czechia
India
Hungary
Philippines
Afghanistan
Honduras
Japan
D Republic
Sweden
Iraq
Puerto Rico
Israel
Turkey
Egypt
Bahrain
Panama
Saudi Arabia
Nepal
Costa Rica

**(c)**

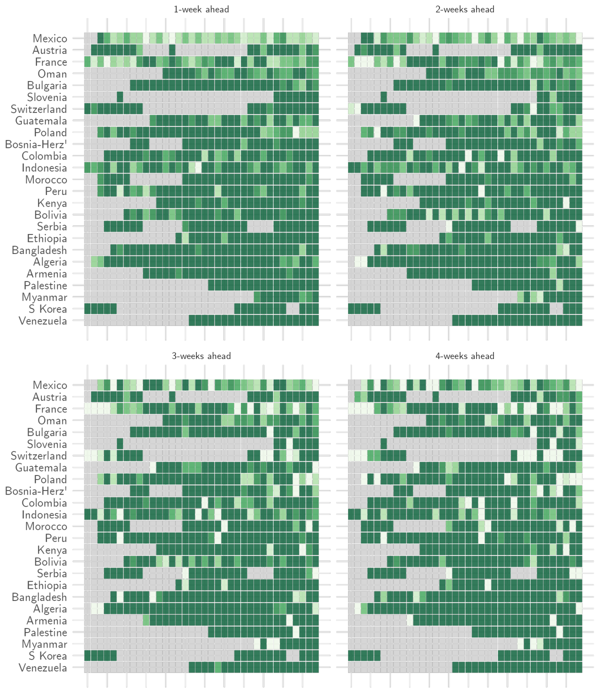

**(d)**

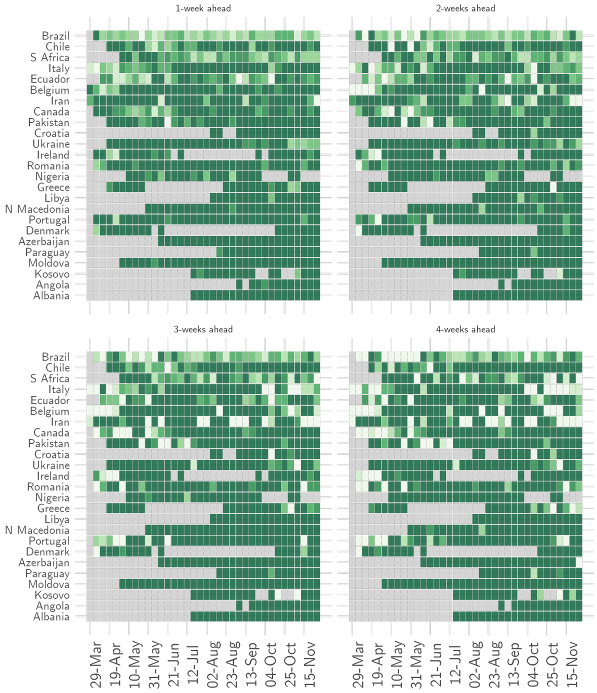

**Figure 12.** The proportion of observations in the 95% CrI of the forecasts for 1-week, 2-week, 3-week, and 4-week ahead for each week of forecast (x-axis) and for each country (y-axis). Panels (a)-(d) present results for all countries included in the analysis. Gray cells indicate weeks where a country was not included in the analysis because the number of deaths did not meet the threshold (SI Sec. 2).

#### Medium-term phase

##### Misclassified epidemic phase

| | Phase using $R^S$ | | | | |
|---|---|---|---|---|---|
| Phase using $R^{curr}$ | Definitely decreasing | Likely decreasing | Definitely growing | Likely growing | Indeterminate |
| Definitely decreasing | 0.00% (0) | 100.00% (253) | 0.00% (0) | 0.00% (0) | 0.00% (0) |
| Likely decreasing | 72.73% (328) | 0.00% (0) | 0.00% (0) | 0.00% (0) | 27.27% (123) |
| Definitely growing | 0.00% (0) | 0.00% (0) | 0.00% (0) | 56.29% (1513) | 43.71% (1175) |
| Likely growing | 0.00% (0) | 0.00% (0) | 0.92% (30) | 0.00% (0) | 99.08% (3239) |
| Indeterminate | 1.68% (31) | 79.35% (1460) | 0.00% (0) | 18.97% (349) | 0.00% (0) |

**Table 4.** In country-days where the phase definitions using $R_t^{curr}$ (shown along rows) and $R_t^S$ (shown along columns) were different, $R_t^S$ most frequently mis-classified the phase as a phase with greater uncertainty. The numbers in parenthesis indicate the number of country-days for a given combination of phase in row and column.

### 7 Code

All analysis was carried out in R version 4.0.2. The code for the analysis is available as orderly [9] project at `https://github.com/mrc-ide/covid19-forecasts-orderly`. DeCa model is available as an R package at `https://github.com/sangeetabhatia03/ascertainr`. The accompanying R package `https://github.com/mrc-ide/rincewind` contains utility functions for creating the figures and processing model outputs.

covid19.who.int.
